# Tryptophan metabolites are associated with gut-brain alterations in functional dyspepsia

**DOI:** 10.1101/2025.11.23.25340799

**Authors:** Matthias Ceulemans, Lucas Wauters, Nathalie Weltens, Greet Vandermeulen, Henriette de Loor, Pieter Evenepoel, Raul Y. Tito, Arnau Vich Vila, Muriel Derrien, Boushra Dalile, Jan Tack, Kristin Verbeke, Jeroen Raes, Lukas Van Oudenhove, Tim Vanuytsel

## Abstract

**Background:** Disturbances in the microbiota-gut-brain axis are thought to contribute to the pathophysiology of disorders of gut-brain interaction, including functional dyspepsia (FD), although comprehensive human data remain scarce.

**Objective:** We aimed to study the relationships among microbiota-produced metabolites including tryptophan metabolites and short-chain fatty acids (SCFA), and functional brain connectivity in FD, in relation to symptomatology.

**Methods:** In 46 patients with Rome IV-diagnosed FD and 30 healthy controls (HC), targeted metabolomics using chromatography and mass spectrometry was conducted to quantify metabolites in blood, urine, and stool. Associations with gut microbiota and symptomatology were tested using fecal quantitative microbiota profiling and validated symptom questionnaires. Resting-state functional magnetic resonance imaging in 27 patients and 36 HC enabled analysis of functional connectivity in selected brain networks.

**Results:** Patients with FD exhibited distinct profiles of tryptophan metabolites and SCFA with higher urinary indole-3-acetate (IAA, *P*=0.018), lower serum kynurenine (*P*=0.030) and lower plasma propionate (*P*=0.0055) concentrations. FD-specific metabolite alterations were associated with more severe GI and psychological symptoms. The fecal microbiota profile was similar between FD and HC. Complementary analyses demonstrated significant alterations in resting-state brain connectivity of 44 predefined regions between FD and HC, while a connectivity-based classifier discriminated FD from HC (82.3% sensitivity, 66.7% specificity, *P*<0.0001). Differences in connectivity measures mediated the higher urinary IAA levels in FD.

**Conclusion:** Dysregulated functional brain connectivity supports an objective pathophysiology in FD. Alterations in specific tryptophan metabolite and SCFA levels were linked to symptomatology, highlighting their potential as biomarkers, and warranting further investigation on microbiota modulating therapies for FD.

**ClinicalTrials.gov:** www.clinicaltrials.gov NCT03545243

**Key Summary:** Summarise the established knowledge on this subject.

○ Functional dyspepsia (FD) is a prevalent disorder of gut-brain interaction (DBGI), frequently associated with psychological comorbidities.
○ The microbial tryptophan metabolite indole-3-acetate (IAA) relays psychological stress to the gut in mice.

What are the significant and/or new findings of this study?

○ Patients with FD have a dysregulated metabolite profile characterized by elevated urinary IAA, and lower serum kynurenine and plasma propionate.
○ Metabolite alterations are linked to comorbid psychological symptoms in FD in the absence of major gut microbiota shifts.
○ Altered functional brain connectivity discriminates FD from healthy controls and mediates elevated urinary IAA levels.

## Introduction

Functional dyspepsia (FD) is a chronic condition characterized by specific meal-related and painful symptoms perceived in the gastroduodenal region. Diagnosed by the Rome IV criteria in the absence of any organic or structural disease, hallmark FD symptoms include postprandial fulness, early satiation, epigastric pain and burning^1^. With an estimated worldwide prevalence of over 7%, FD affects considerably more individuals than irritable bowel syndrome (IBS)^2^, despite more limited public awareness in comparison. Both functional gastrointestinal (GI) disorders were recently relabeled as disorders of gut-brain interaction (DGBI)^3^, to better reflect their pathophysiology, presumed to involve dysfunction of intricate gut-brain interplay. Besides an impaired quality of life^4^ and common overlap with psychological distress^5^, FD imposes a substantial socio-economic burden due to repeated healthcare visits and a lack of cost-effective treatment options^6^, highlighting the pressing need for increased insight in its pathophysiology to support clinical diagnosis and guide therapy selection.

Research on pathophysiological mechanisms underlying FD symptoms has primarily focused on mucosal immune activation^7^, epithelial barrier defects^8^ and gut microbial alterations^9^. Specific microbial shifts were found in the proximal GI tract^10^, with most evidence for duodenal microbial changes, including increased abundance of *Streptococcus*^10^ and reduced *Firmicutes*^11^, *Neisseria*^12^ and *Porphyromonas*^12^ abundance in FD compared to controls. An independent association between the onset of FD and psychological disorders such as anxiety and mood disorders suggests an important role of gut-brain communication^13^. This gut-brain crosstalk can potentially be influenced by the gut microbiota. Important metabolites produced by gut microbes, most notably short-chain fatty acids (SCFA) and aromatic amino acid (AAA; tryptophan, phenylalanine and tyrosine) metabolites, are thought to directly or indirectly play a role in various homeostatic processes including microbiota-gut-brain communication^14, 15^. Indole-3-acetate (IAA), a gut microbiota-derived tryptophan metabolite, has been shown to relay psychological stress to the gut in mice^16^, offering an attractive hypothesis for the high psychological symptom burden in FD and related DGBI. However, comprehensive data on the role of microbial metabolites in altered microbiota-gut-brain crosstalk in FD are currently lacking.

To address this knowledge gap, we performed targeted metabolomic and fecal microbiota profiling in a prospectively recruited cohort of patients with FD and matched healthy controls (HC). We combined extensive symptom profiling and functional magnetic resonance imaging (fMRI) of the brain to evaluate central alterations related to FD pathophysiology. Lastly, using regression, classification and mediation models, we identified a link between urinary IAA, GI and psychological symptom profiles, and functional brain alterations in FD.

## Materials and methods

### Study design and participants

Patients with Rome IV-defined FD (n=46) and matched healthy controls (HC) (n=30) were prospectively recruited in a single center to undergo sampling of blood, urine and feces for extensive metabolite and microbiota profiling. In- and exclusion criteria are listed in the **Suppl.Methods**. During the study visit, participants completed validated questionnaires assessing GI and extraintestinal symptoms, as well as dietary intake (**Suppl.Methods**). The study was approved by the University Hospitals / KU Leuven Ethics Committee for Research (S60953/S60984) and registered at ClinicalTrials.gov (NCT03545243). All participants provided written informed consent before the start of the study. The study was conducted in accordance with the Declaration of Helsinki and the Guidelines for Good Clinical Practice. All co-authors had access to the study data and have reviewed and approved the final manuscript.

### Sample collection

During study visits, qualified staff collected blood samples in the morning (to avoid diurnal variation) in fasted state (≥ 6h) in EDTA and SST tubes (both BD Vacutainer) for plasma and serum collection, respectively. Blood samples were centrifuged, aliquoted, and serum (amino acid metabolite analyses, **Fig.2A**) and plasma (short-chain fatty acids (SCFA) analyses) were stored at -80°C until targeted metabolite analyses (**Suppl.Methods**). Participants provided a fasted spot urine sample, which was stored at -80°C until analysis (**Suppl.Methods**). Within two days of the study visit, participants collected a stool sample for microbiota and SCFA analysis at home in a container provided for this purpose, which was immediately frozen (-18°C) at participants’ own freezer and transported to the lab with ice packs, where samples were stored at -80°C until analysis (**Suppl.Methods**). Participants scored the consistency of their stool sample using the Bristol Stool Scale (BSS) as a proxy for intestinal transit time^17^.

### Functional magnetic resonance imaging

Optional rs-fMRI and T1 structural MRI measurements were performed in consenting individuals (27 patients with FD and 20 HC) at the University Hospitals Leuven Radiology Department using a 3T Philips Achieva DStream MRI scanner (32-channer head coil). The dataset was complemented with rs-fMRI measurements of a previously recruited cohort of 16 additional HC who underwent identical fMRI procedures (same scanner, same head coil, identical acquisition parameters). Processing and analysis of fMRI data was performed in batch and described in the **Suppl.Methods**.

### Statistical analyses

Statistical analyses were performed in R (v4.5.1) and described in detail in the **Suppl.Methods**.

## Results

### High psychological symptom burden in FD

To investigate the contribution of gut microbial metabolites to the pathophysiology of FD, we studied a well-characterized cohort of 46 patients with Rome IV-diagnosed FD and 30 HC. The absence of endoscopic alterations was confirmed during esophagogastroduodenoscopy in all participants. The characteristics of this cohort have been described before^18^ (**Suppl.Table1**). Briefly, patients and controls were matched for age, sex, body mass index (BMI), and geographic origin (all *P*>0.1). Thirty nine percent of patients with FD (n=18/46) were taking PPI at the time of sampling, while none of the HC were taking PPI. Besides more severe GI-specific symptoms (LPDS and PAGI-SYM; both *P<*0.0001), patients with FD presented with significantly higher symptom scores for anxiety (GAD7), perceived stress (PSS), depression (PHQ9), extraintestinal symptoms (PHQ12), GI-specific anxiety (VSI) and posttraumatic stress (PTSD) (all *P<*0.0004) (**Fig.1**, **Suppl.Table2**, **Suppl.Results**) as described before^5^.

**Figure 1:**
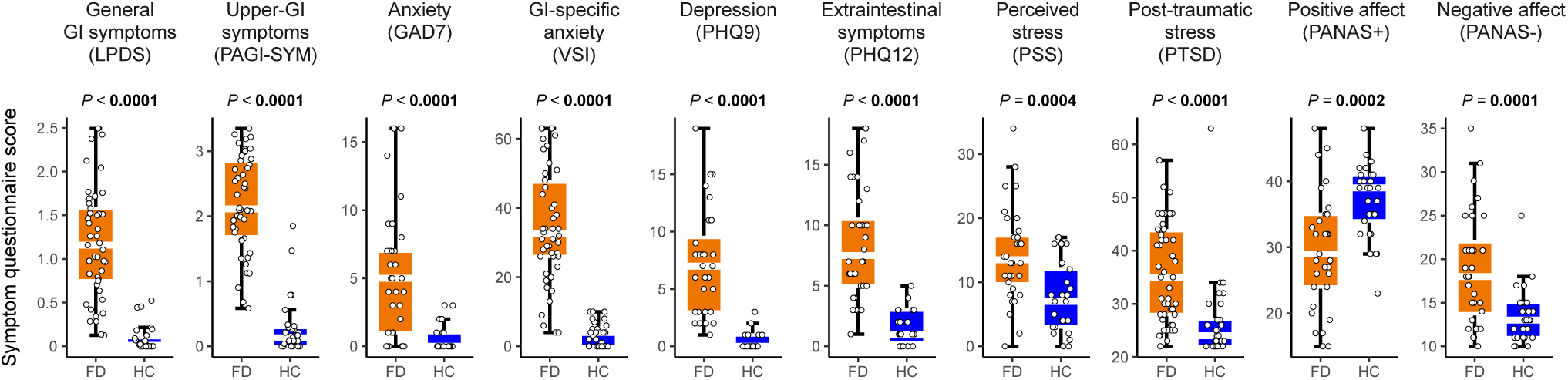
High comorbid psychological symptom burden in FD. Between-group differences in gastrointestinal (LPDS, PAGI-SYM) and psychological (GAD7, VSI, PSQ9, PHQ12, PSS, PTSD, PANAS+, PANAS-) symptoms. Linear regression models with ‘PPI intake’ nested in the grouping variable. *P* values represent main group effects.

### Patients with FD exhibit a distinct metabolite profile

We measured the concentrations of a relevant selection of host and microbiota-derived aromatic amino acids (AAA) and key AAA metabolites (**Fig.2A**) using targeted ultra-performance liquid chromatography-tandem mass spectrometry. Principal component analysis (PCA) and permutational analysis of variance (PERMANOVA) revealed a significantly distinct serum-derived metabolic signature between FD and HC (R^2^=0.031, *P_PERMANOVA_=*0.012) (**Fig.2B**), driven by a significantly different serum microbial AAA metabolite signature (R^2^=0.033, *P_PERMANOVA_=*0.030) (**Suppl.Fig.1A**). The urinary AAA metabolite profile was not different between both groups (R^2^=0.024, *P_PERMANOVA_=*0.11) (**Suppl.Fig.1B**), although the profile of urinary microbial AAA metabolites differed significantly between FD and HC (R^2^=0.037, *P_PERMANOVA_=*0.040) (**Fig.2C**).

**Figure 2:**
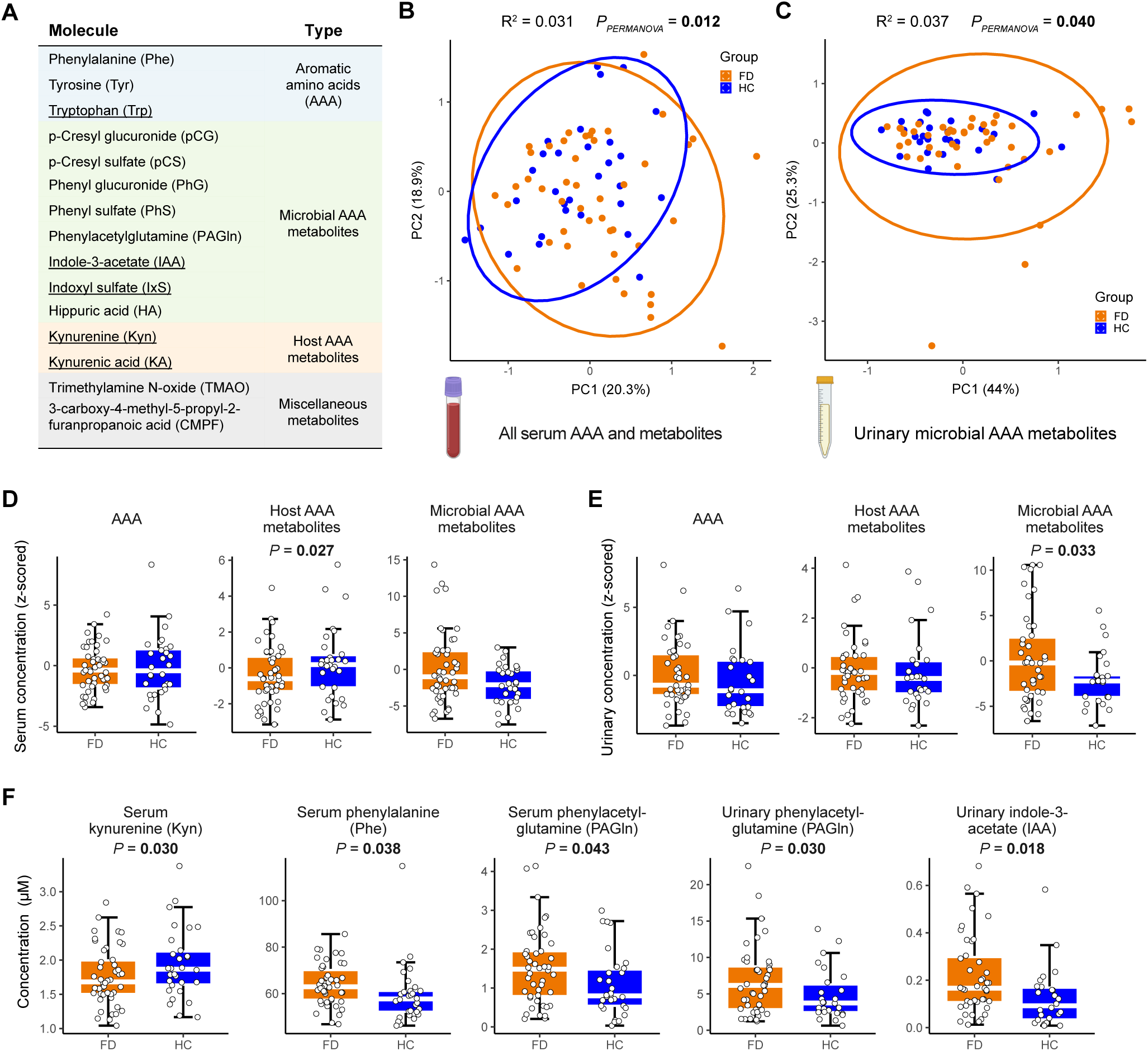
Patients with FD exhibit a distinct amino acid metabolite profile. (**A**) Aromatic amino acids (AAA) and AAA metabolites included in the targeted metabolomics approach. Tryptophan and its metabolites are underlined. Principal component analysis of (**B**) serum AAA and AAA metabolites, and (**C**) urinary microbial AAA metabolites. Permutational analysis of variance (PERMANOVA, 10,000 permutations) quantified explained proportion of variance (R^2^) by ‘Group’. Between-group differences in standardized concentrations of AAA and AAA metabolites aggregated per origin, measured in (**D**) serum and (**E**) urine, (**F**) and levels of selected individual AAA metabolites. Linear regression with main ‘Group’ effect and ‘PPI intake’ nested in the grouping variable, adjusted for protein intake. *P* values represent main ‘Group’ effects (**D**-**F**).

Next, the concentrations of analyzed molecules were standardized and aggregated by their origin, i.e. AAA, host-derived AAA metabolites and microbial AAA metabolites (**Fig.2A**). Serum (η_p_^2^=0.10, *P*=0.027) but not urinary host AAA metabolites (η_p_^2^=0.00004, *P*=0.97) were lower in FD (**Fig.2D-E**), after adjusting for protein and PPI intake. In line with PCA results, urinary microbial AAA metabolites (η_p_^2^=0.11, *P*=0.033) were elevated in FD (**Fig.2E**). Systemic and urinary AAA levels were similar between groups (all *P*>0.1) (**Fig.2D-E**). On the individual metabolite level, serum host-derived AAA metabolite alterations were driven by lower serum kynurenine (Kyn) (η_p_^2^=0.10, *P*=0.030), while urinary phenylacetylglutamine (PAGln) (η_p_^2^=0.12, *P*=0.030) and indole-3-acetate (IAA) (η_p_^2^=0.14, *P*=0.018) were elevated in FD (**Fig.2F**). Serum phenylalanine (η_p_^2^=0.09, *P*=0.038) and PAGln (η_p_^2^=0.09, *P*=0.043) levels were higher in FD (**Fig.2F**). Tryptophan and tyrosine levels in serum or urine were not different between groups.

Fecal and plasma acetate, butyrate and propionate were analyzed using gas chromatography-mass spectrometry. Concentrations of all measured individual and total SCFA in both feces and plasma were lower in FD compared to HC (**Fig.3A-B**), with systemic propionate (η_p_^2^=0.16, *P*=0.0055) and total SCFA (η_p_^2^=0.17, *P*=0.0044) reaching statistical significance, independent of PPI and fiber intake, the major substrate for SCFA production (**Fig.3B**).

**Figure 3:**
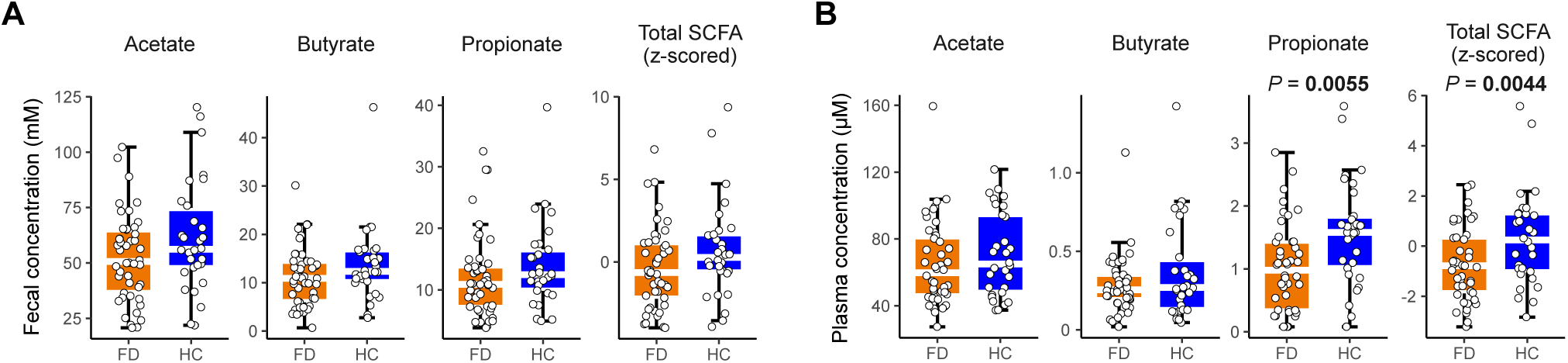
Reduced plasma propionate levels in patients with FD. Between-group differences in concentrations of individual and total short-chain fatty acids (SCFA) measured in (**A**) feces and (**B**) plasma. Linear regression with main ‘Group’ effect and ‘PPI intake’ nested in the grouping variable, adjusted for fiber intake. *P* values represent main ‘Group’ effects.

### IAA, Kyn and propionate are associated with gastrointestinal and comorbid psychological symptoms in FD

Gut microbiota-derived IAA was recently shown to relay psychological stress to the gut in mice, while increased IAA levels in patients with major depressive disorder demonstrated its relevance in a human context^16^. To explore the clinical relevance of IAA and other dysregulated metabolites in FD, we performed correlation analyses between symptom outcomes and metabolites. Consistently, urinary IAA, plasma propionate and serum Kyn were among the metabolites most strongly associated with symptomatology (**Fig.4A**, **Suppl.Table4**). To confirm these exploratory correlations, we constructed linear regression models to assess associations between metabolites and participants’ GI and comorbid psychological symptoms. Urinary IAA was associated with general anxiety, depression, extraintestinal and GI-specific symptoms (**Suppl.Fig.2A**, **Suppl.Table5**), confirming the previously suggested link between IAA and psychological distress^16^. Plasma propionate levels were significantly and inversely associated with symptom levels except for perceived stress (**Suppl.Fig.2B**, **Suppl.Table5**). Similarly, serum Kyn, a tryptophan metabolite of non-microbial origin, was inversely associated with symptoms except for GI-specific anxiety (**Suppl.Figure2C**, **Suppl.Table5**). Medium to large effect sizes of these metabolite-symptom associations persisted when controlling for dietary protein (IAA, Kyn) or fiber (propionate) and PPI intake (**Suppl.Table6**), suggesting independent associations.

**Figure 4:**
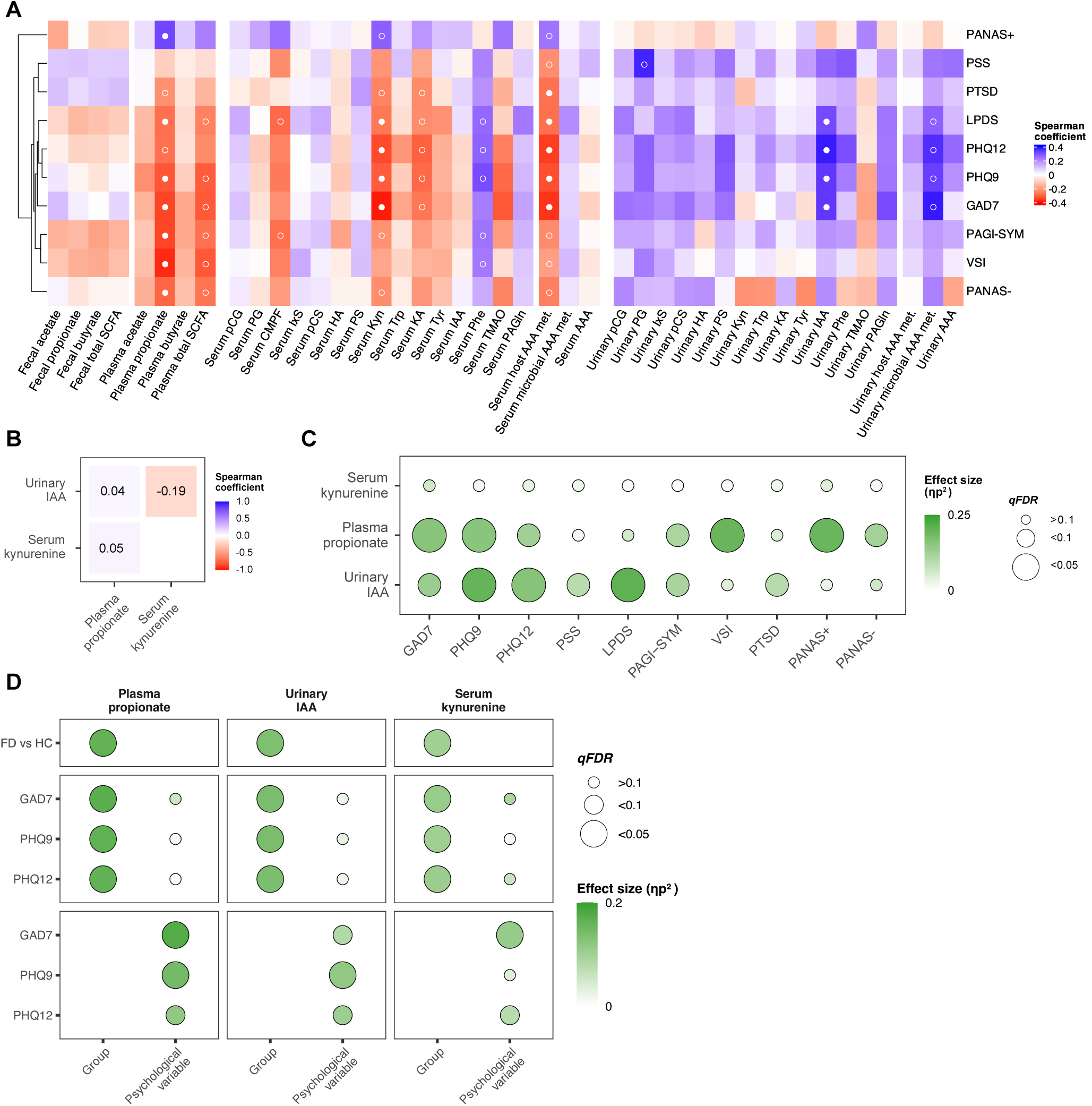
IAA, propionate and Kyn are associated with gastrointestinal and comorbid psychological symptoms in FD. (**A**) Heatmap of associations between symptom scores and metabolites. Color scale depicts Spearman correlation coefficients. Benjamini-Hochberg adjusted significance depicted as open (*q_FDR_*<0.1) or filled circles (*q_FDR_*<0.05). (**B**) Heatmap of pairwise correlations between significantly altered metabolites of interest in FD. Color scale and values depict Spearman correlation coefficients. (**C**) Multiple linear regression analysis between significantly altered metabolites of interest in FD and symptom scores. (**D**) Linear regression analyses between metabolites of interest with FD (top panels) or comorbid psychological variables separately (lower panels), and with FD adjusted for symptom variables (middle panels). All models were corrected for fiber and protein intake where appropriate as well as PPI intake. Circle sizes correspond to Benjamini-Hochberg adjusted *P* values (*q_FDR_*). Color scale depicts effect sizes (η_p_^2^).

Of note, urinary IAA, plasma propionate and serum Kyn were only minimally correlated with each other (**Fig.4B**). To assess the individual contribution of each metabolite of interest on the different symptom variables, we used multiple linear regression analysis with correction for fiber and protein intake. While urinary IAA was strongly associated with depression, extraintestinal and general GI symptoms, the association between plasma propionate with upper-GI symptoms, general and GI-specific anxiety, and both positive and negative affect had the highest effect sizes. In contrast, serum Kyn only contributed minimally to each of the metabolite-symptom associations (**Fig.4C**, **Suppl.Table7**). Individual linear models confirmed the strong association of plasma propionate, urinary IAA and serum Kyn with both FD, as well as with comorbid psychological symptoms. However, when adjusting for psychological symptoms, only the case-control effect on these metabolites persisted, indicating a higher relative contribution of FD over psychological comorbidities to the metabolite-symptom associations (**Fig.4D**, **Suppl.Table8**).

To investigate whether the shifts in specific metabolites were linked to alterations in the gut microbiota, we performed quantitative microbiota profiling (QMP) on fecal samples^19^. Despite similar stool consistency and microbial density, diversity or composition between FD and HC (**Fig.5A-E**), the quantitative abundance of select taxa (decreased *Odoribacter*, *Turicibacter* and unclassified *Clostridia*; increased *Flavonifractor*; all *q_FDR_*=0.075) tended to differ in patients with FD, independent of PPI intake and intestinal transit time (**Fig.5F**, **Suppl.Results**). In addition, the fecal genera associated with metabolic changes and psychological symptom profiles clustered separately, suggesting the absence of a shared microbial signature underlying these alterations in FD (**Suppl.Fig.4**, **Suppl.Results**).

**Figure 5:**
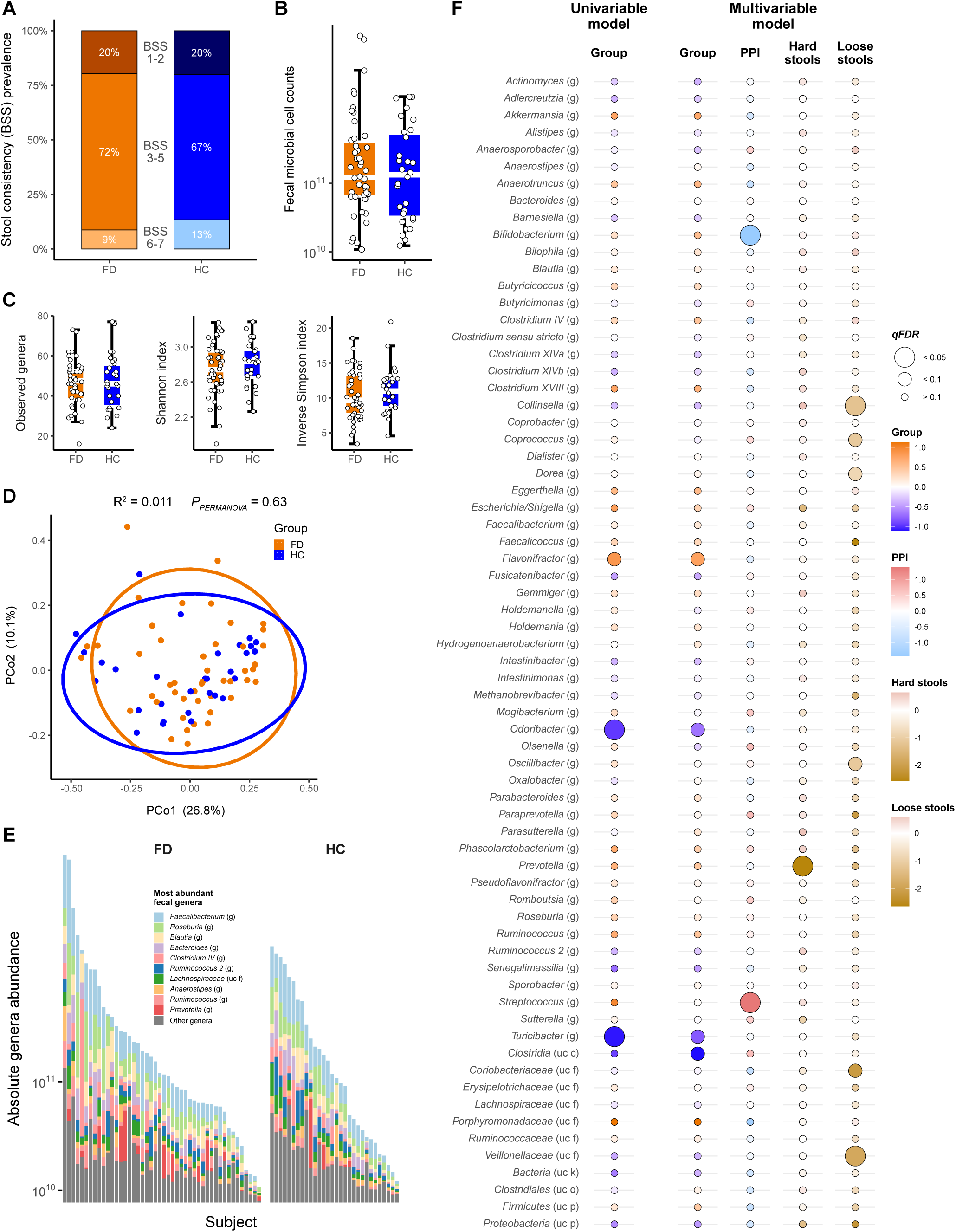
The fecal microbiota in FD is similar compared to HC and shaped by intestinal transit time. (**A**) Distribution of stool sample consistency types between FD and HC (Bristol Stool Scale, BSS). (**B**) Fecal microbial load (flow cytometric microbial cell counts) comparison between groups (left) or BSS category (right). (**C**) Differences in fecal microbial alpha diversity indices (genus-level) between groups. (**D**) Principal coordinate analysis (PCoA, Bray-Curtis) of fecal microbiota in FD and HC. Permutational analysis of variance (PERMANOVA, 10,000 permutations) quantified explained proportion of variance (R^2^) by ‘Group’. (**E**) Absolute abundance of ten most abundant fecal bacterial genera. (**F**) Univariable (between-group differences) and multivariable (between-group differences controlled for proton pump inhibitor (PPI) intake and intestinal transit time (BSS)) fecal microbial genera abundance analysis. Circle sizes correspond to Benjamini-Hochberg adjusted *P* values (*q_FDR_*). Color scale depicts individual effect sizes.

### Resting-state functional connectivity in salience, emotional-arousal, and central autonomic brain networks is altered in FD and classifies FD and HC with reasonable accuracy

Studies that comprehensively consider gut microbiota metabolites and brain components are scarce. Therefore, participants in this study underwent optional resting-state functional magnetic resonance imaging (rs-fMRI) of the brain, completed by 27 patients with FD and 20 HC, supplemented with data from 16 additional HC who underwent identical rs-fMRI procedures. This cohort of 36 HC had a similar sex distribution compared to the 27 FD patients (25 females (69%) versus 22 females (81%), *P*=0.38) but was younger on average (mean age (years) ± standard deviation age: 28±9 years vs 34±12, *P*=0.015).

Multivariate general linear model (GLM) analyses of a region-of-interest (ROI)-to-ROI connectivity matrix incorporating 44 a priori selected ROIs involved in salience, emotional-arousal and central autonomic networks^20^ (**Suppl.Table9**), demonstrated a significant difference in connectivity between FD and HC for the right inferior frontal gyrus - pars triangularis (ventrolateral prefrontal cortex, vlPFC) (ROI mass=61.27, *P*=0.0013, *P_FWE_*=0.046). This was driven by significantly stronger connectivity between this region with the bilateral rostral anterior cingulate cortex and gyrus rectus, as well as with left medial fronto-orbital gyrus and superior frontal gyrus in FD versus HC (**Fig.6A**, **Suppl.Table10**).

**Figure 6:**
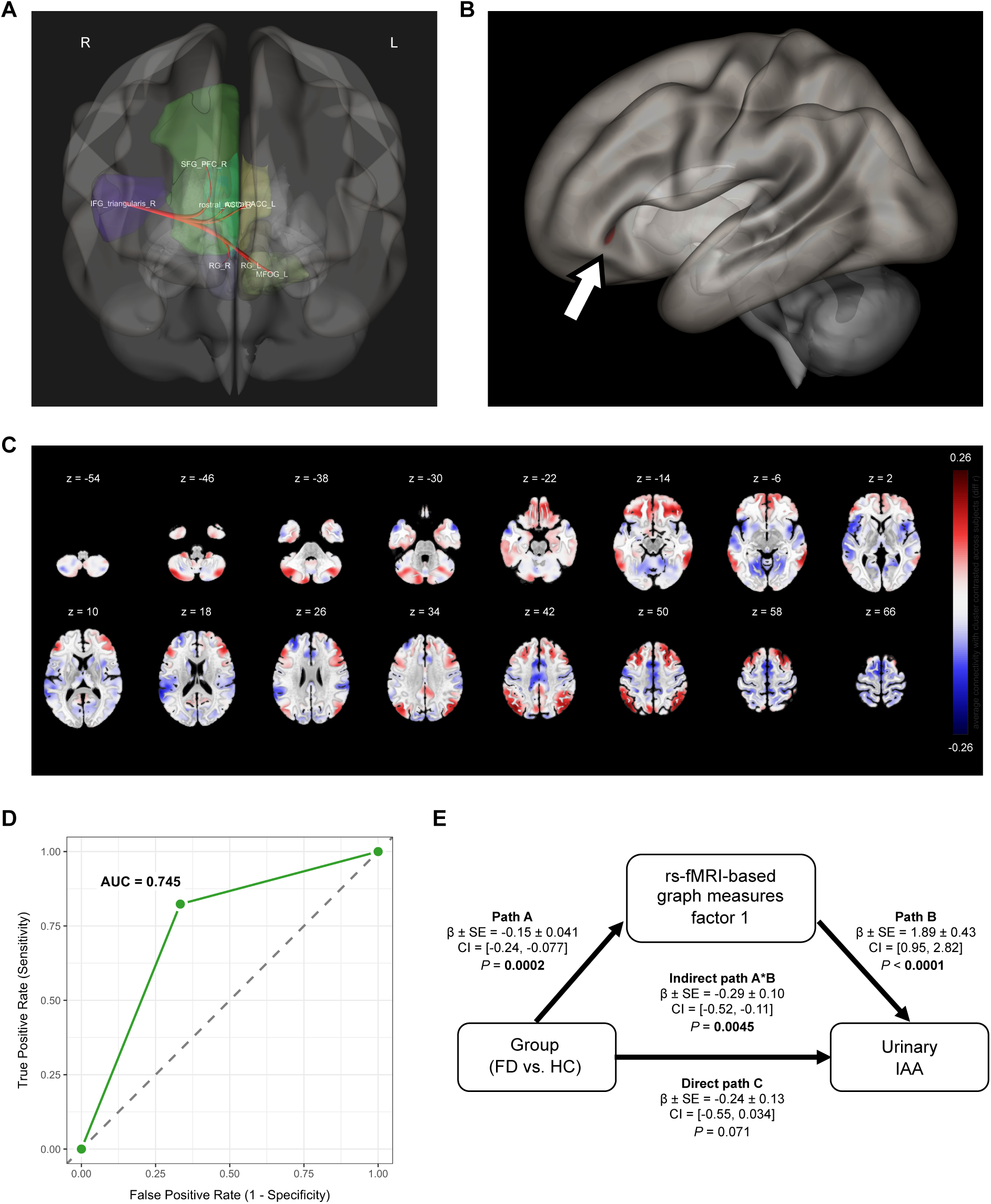
Resting-state functional brain connectivity is altered in FD and mediates elevated urinary IAA levels. (**A**) Between-group difference in region-of-interest (ROI)-to-ROI-based connectivity for the right inferior frontal gyrus (IFG) – pars triangularis (ventrolateral prefrontal cortex, vlPFC). (**B**) Between-group difference in whole-brain voxel-based connectivity for the left vlPFC (arrow). (**C**) Difference in connectivity pattern of the left vlPFC cluster with other brain regions between FD (red) and HC (blue). (**D**) Receiver operating characteristic (ROC) curve and area under the curve (AUC) depicting the performance of brain connectivity network partial least squares discriminant analysis (PLS-DA) classification between patients with FD and HC. (**E**) Mediation of between-group differences on urinary indole-3-acetate (IAA) levels. Significant between-group effect on brain connectivity (“Path A”) and significant association of brain connectivity with urinary IAA (“Path B”). Introducing brain connectivity (resting-state functional magnetic resonance imaging (fMRI)-based graph measures factor 1) as mediator of the between-group effect on urinary IAA reduced the significance of the direct between-group effect on urinary IAA (“Path C”), while the indirect through the mediator (“Path A*B”) remained significant. Model outcomes of the individual mediation paths are summarized as estimate (β) ± standard error (SE) with corresponding bootstrapped 95% confidence intervals (CI).

Next, we applied functional connectivity multivariate pattern analysis (fc-MVPA) as a whole-brain data-driven approach to identify regions in which connectivity with the rest of the brain differs between FD and HC. A significant between-group difference in whole-brain voxel-based connectivity was found for a cluster in the left vlPFC including frontal orbital cortex and inferior frontal gyrus - pars triangularis (peak MNI coordinates [x,y,z]=[-41,33,-7]; k=61 voxels; TFCE=431.76; *P*=0.001; *P_FWE_=*0.048) (**Fig.6B**). This difference was driven by stronger connectivity between this cluster and other (pre)frontal, inferior temporal, and cerebellar areas in FD patients (red), and stronger connectivity between this cluster and insular and parietal regions in HC (blue) (**Fig.6C**).

To test the collective ability of the calculated graph measures to discriminate between patients with FD and controls, they were entered as independent variables in partial least squares discriminant analysis (PLS-DA). PLS-DA with all 540 graph measures as X-variables and group (FD versus HC) as Y-variable was performed and 5-fold cross-validation suggested a 1-factor solution with 55% of X-variables (296/540) exceeding the variable importance plot (VIP)-threshold of 0.8 (van der Voet T²=0, Q²=0.18) (**Suppl.Fig.5A**). The model cumulatively explained 25.3% and 33.3 % of the variance for the X-variables and the Y-variable, respectively (**Suppl.Table12**). Classification performance was adequate and significantly different from chance (75.5% accuracy, 82.3% sensitivity, 66.7% specificity, AUC=0.745, *P*<0.0001) (**Fig.6D**).

### Altered resting-state functional brain connectivity mediates elevated urinary IAA levels in FD

Partial least squares (PLS)-regression with all 540 graph measures as X-variables and urinary IAA as Y-variable was performed and leave-one-out cross-validation suggested a 2-factor solution with 59% of X-variables (320/540) exceeding the VIP-threshold of 0.8 (van der Voet T²=0, Q²=0.04) (**Suppl.Fig.5B**). The model cumulatively explained 26.8% and 81.5% of the variance for the X-variables and the Y-variable, respectively (**Suppl.Table13**). Moreover, the PLS-regression model significantly predicted urinary IAA levels, explaining 81.1% of the variance (Pearson r=0.90, *P<*0.0001) (**Suppl.Fig.5C**). In similar models with plasma propionate or serum Kyn as the Y-variable, the cross-validation procedure did not suggest any factors to retain (based on a minimum value for the root mean PRESS statistic for the null model), indicating that the X-variables (graph measures) did not explain any variance in the Y-variables (**Suppl.Fig.5D**).

Of note, one-way ANOVA revealed that the standardized first factor of the PLS-regression model – explaining 14.9% of the variance in the graph measures – was significantly higher in FD (Cohen’s d=1.13, *P*=0.0008), while the second factor explaining 11.9% of the variance was not different between groups (d=0.29, *P*=0.42) (**Suppl.Fig.5E**). This prompted us to test whether the group difference in urinary IAA levels was mediated by differences in graph measures. For this purpose, we fitted a mediation model with group as the independent (X)-variable, the standardized first graph measures factor of the PLS-regression model predicting urinary IAA as the mediator (M)-variable, and the standardized urinary IAA levels as the dependent (Y)-variable. The indirect effect of group on IAA through the graph measures factor (path A*B) was significant (β±SE=-0.29±0.10; 95% bootstrapped CI=[-0.52,-0.11], *P*=0.0045) while the direct effect of group on IAA (path C) was non-significant (-0.24±0.13, [- 0.55,0.034], *P*=0.071) (**Fig.6E**). The total effect of group on IAA was significant (- 0.53±0.14, [-0.82,-0.26], *P*=0.0001), confirming that differences in brain-connectivity measures mediated the higher urinary IAA levels in FD.

## Discussion

The bidirectional interplay between the gut microbiota and the central nervous system, known as the microbiota-gut-brain axis, is well established through preclinical and clinical studies^21–23^. Nevertheless, its role in DGBI including FD remains incompletely understood. Here, we provide novel evidence for dysregulated SCFA and tryptophan metabolism in FD in the absence of major microbial alterations, linked to a worsened symptom profile and altered functional brain connectivity.

We identified a dysregulated metabolite signature in FD compared to HC, with a marked reduction in circulating kynurenine but increased urinary IAA levels. This suggests a shift in tryptophan metabolism from the host kynurenine pathway toward microbial indole production. Interestingly, a decrease in serum kynurenine levels has been described in healthy men after acute stress induction^24^, although indole metabolites and the microbiota were not evaluated. Furthermore, *Lactobacillus murinus*-produced IAA has recently been demonstrated to relay psychological stress to the gut in mice, where it caused impaired stem cell differentiation. In patients with major depressive disorder (MDD), *Lactobacillus* abundance and fecal IAA levels were higher compared to controls^16^. In contrast, supplementation of IAA partially alleviated colitis in mice by restoring intestinal barrier function^25^, suggesting differential effects of IAA depending on the inflammatory and genetic context or mouse model.

All major SCFA concentrations (acetate, butyrate and propionate) measured in the circulation and feces were numerically lower in FD compared to controls, with the reduction in systemic propionate levels being most pronounced and statistically significant. To the best of our knowledge, this is the first study to demonstrate a reduction in SCFA levels in FD compared to HC. This finding is especially interesting given that SCFA – important products of microbial carbohydrate degradation – can influence central processes when reaching the systemic circulation^14^. Lower systemic SCFA levels are in line with recent findings of decreased circulating butyrate and propionate levels in patients with MDD^26^, suggesting that limited SCFA availability is particularly linked with the central and psychological aspects of FD.

Importantly, altered metabolite levels in FD correlated with patient-reported GI and psychological symptoms. However, we observed a higher relative contribution of FD diagnosis over individual symptoms, indicating a specific association with psychological comorbidities tied to FD as opposed to psychological disorders alone. This underscores the clinical relevance of metabolite dysregulation in DGBI with a concomitant psychopathology, although cause and consequence cannot be untangled from our cross-sectional analysis.

Despite significant shifts in metabolite profiles, we found no major change in microbial load and diversity of the fecal microbiota in FD. Previous studies on microbial alterations in FD focused on the gastroduodenal environment, consistently demonstrating increased abundance of typical upper-GI bacteria such as *Streptococcus*^9, 10^. A recent case-control study from South-Korea in 12 patients with FD found an overall similar fecal microbiota composition compared to 16 controls as assessed by alpha and beta diversity^27^, which we corroborated in a larger cohort with a different geographical origin. We and others reported on the beneficial effects of probiotics for FD in randomized placebo-controlled trials, associated with proliferation of SCFA-producing microbes^28, 29^. This warrants further investigation on the anticipated beneficial effects of intervention with probiotics, as well as prebiotics, aimed at restoring circulating SCFA levels in randomized controlled trials.

One of the most intriguing findings of our study is the significantly altered brain connectivity in patients with FD compared to HC, which objectively validates FD as a disorder of gut-brain interaction. Altered connectivity of the ventrolateral prefrontal cortex – a region implicated in reward valuation and pain modulation – with other emotional-arousal, salience, executive, and central autonomic network regions was found to be a unique brain feature in FD. Moreover, assessment of the global organization of brain connectivity networks through application of graph theoretical analysis pointed toward an overall altered network topology between FD and HC. This was further substantiated by an accurate classification of patients and controls based on this topological analysis. These findings corroborate the results from multiple studies that reported on functional brain alterations in FD (reviewed in^30–33^), supporting the concept that the pathogenesis of this disorder likely involves a combination of central and peripheral impairments. However, this study is the first to integrate brain connectivity measurements with metabolite levels in a translational hypothesis-driven fashion. Indeed, the graph measures partial least squares regression model explained a substantial portion of the variation in urinary IAA levels. Although causality cannot be inferred from this cross-sectional study, statistical mediation analysis suggested that elevated IAA might be driven by the observed brain connectivity changes in FD. In addition, this translates the preclinical findings by Wei *et al*. of increased IAA levels in various mouse models of psychological stress^16^, to a human context of a disorder where psychological stress is thought to play an important role.

Some limitations inherent to the design of the study should be acknowledged. This study applied targeted metabolomics, measuring a limited number of AAA and their metabolites, leaving other potentially relevant metabolites unexplored, but ensuring an optimized and validated analysis of several key tryptophan metabolites and major SCFA in multiple human samples. Our microbial analysis was conducted by 16S rRNA gene sequencing. Future studies using higher resolution metagenomic sequencing should focus on identifying the pathways underlying these observed metabolomic alterations. In the current study, fMRI was restricted to resting state measurements, whereas task-based imaging could identify alterations in connectivity and activity in specific disorder-related contexts including stress, anxiety or pain. Lastly, findings from this relatively small single-center study should be confirmed in larger and diverse populations, controlling for the effects of geography and diet. Nevertheless, current metabolite analyses were adjusted for major dietary confounders including protein and fiber intake, as assessed by validated food frequency questionnaires, as well as PPI intake – a prevalent and important confounder.

Taken together, our study is the first to integrate peripheral microbiota-related alterations with central functionality in FD, adding to the growing body of evidence for a disturbed microbiota-gut-brain axis in FD. The first human data on alterations in tryptophan and short-chain fatty metabolism in FD – linked to its typical symptomatology – suggest they can be actionable targets in this difficult-to-treat disorder to restore microbiota-gut-brain homeostasis by dietary, probiotic or prebiotic intervention, which deserves continued investigation.

## Supporting information

Supplementary methods and results

## Supplementary Figure legends

**Supplementary Figure 1: Patients with FD exhibit a distinct serum microbial metabolite profile.** Principal component analysis of (**A**) serum microbial AAA metabolites, and (**B**) urinary AAA and AAA metabolites. Permutational analysis of variance (PERMANOVA, 10,000 permutations) quantified explained proportion of variance (R^2^) by ‘Group’.

**Supplementary Figure 2: IAA, propionate and Kyn are associated with gastrointestinal and comorbid psychological symptoms in FD.** Linear relationships between psychological (GAD7, PSQ9, PHQ12) and gastrointestinal (LPDS) symptom scores with (**B**) urinary indole-3-acetate (IAA), (**C**) plasma propionate, and (**D**) serum kynurenine (Kyn).

**Supplementary Figure 3: Stool consistency is associated with fecal microbiota composition.** Differences in fecal microbial (**A**) cell counts and (**B**) alpha diversity indices (genus-level) by stool sample consistency. Linear regression with main ‘Group’ effect and ‘PPI intake’ nested in the grouping variable or main ‘BSS category’ effect. *P* values represent main ‘BSS category’ effects. (**C**) Principal coordinate analysis (PCoA, Bray-Curtis) of fecal microbiota samples by stool sample consistency (Bristol Stool Scale, BSS). Permutational analysis of variance (PERMANOVA, 10,000 permutations) quantified explained proportion of variance (R^2^) by ‘BSS category’.

**Supplementary Figure 4: Specific taxa are associated with metabolite and symptom profiles.** Heatmap of associations between altered symptom scores and metabolites in FD with (**A**) differentially abundant fecal genera between FD and HC, and (**B**) twenty most abundant fecal genera. Color scale depicts Spearman correlation coefficients. Benjamini-Hochberg adjusted significance depicted as open (*q_FDR_* < 0.1) or filled circles (*q_FDR_*< 0.05).

**Supplementary Figure 5: Resting-state functional brain connectivity accurately predicts urinary IAA levels.** Factor loading plots for (**A**) partial least squares discriminant analysis (PLS-DA) and (**B**) PLS regression analysis using functional connectivity-based graph measures (X-variables). Plots depict Y variables in blue (**A**: FD and HC, **B**: urinary IAA), individual FD (black circles) and HC (black triangles) subject loadings, and X-variables (green dots, 540 resting-state functional magnetic resonance imaging (fMRI)-based graph measures) to enter the final PLS-DA and PLS regression models, respectively. (**C**) Linear relationship between observed urinary IAA levels and predicted urinary IAA levels by PLS regression. Pearson correlation coefficient (r) and *P* value as indicated. (**D**) Leave-one-out cross-validation procedure outcomes for urinary IAA, plasma propionate and serum kynurenine (Kyn) suggested a 2-factor solution based on a minimum value for the root mean PRESS statistic for the null model for urinary IAA, while 0 factors were suggested for propionate and Kyn. (**E**) Between-group differences in standardized factors of the PLS regression model explaining 14.9% and 11.9% variance in the graph measures, respectively. One-way analysis of variance.

## Supplementary Table legends

**Supplementary Table 1: Overview of study participant characteristics.** Baseline characteristics of patients with functional dyspepsia (FD, n = 46) compared to healthy controls (HC, n = 30), analyzed by unpaired Wilcoxon tests for continuous data and Fisher’s exact tests for categorical data. Means ± standard deviation (SD) or n (%) as indicated. IBS, irritable bowel syndrome.

**Supplementary Table 2: Associations between symptom variables, study group and PPI intake.** Gastrointestinal and comorbid extraintestinal symptoms were assessed by symptom questionnaires. Data were analyzed using linear regression, with inclusion of ‘PPI intake’ nested in the grouping variable (FD vs. HC). Main ‘group’ and ‘PPI’ effects are reported with corresponding effect sizes (partial eta squared values, η_p_^2^). PAGI-SYM, Patient Assessment of Upper GI Symptom Severity; LPDS, Leuven Postprandial Distress Scale; GAD, Generalized Anxiety Disorder; PHQ, Patient Health Questionnaire; VSI, Visceral Sensitivity Index; PTSD, Post-Traumatic Stress Disorder; CTQ, Childhood Trauma Questionnaire; PANAS, Positive and Negative Affect Schedule.

**Supplementary Table 3: Food frequency questionnaire-derived estimation of daily macronutrient intake.** Macronutrient intake was estimated using validated food frequency questionnaires. Data were analyzed using linear regression, with inclusion of ‘PPI intake’ nested in the grouping variable (FD vs. HC). Main ‘group’ and ‘PPI’ effects are reported with corresponding effect sizes (partial eta squared values, η_p_^2^).

**Supplementary Table 4: Correlation analysis between symptom variables and metabolites.** Non-parametric correlations between symptom variables and metabolites were reported as Spearman coefficients (r). SCFA, short-chain fatty acids; AAA, aromatic amino acids; CMPF, 3-carboxy-4-methyl-5-propyl-2-furanpropanoic acid; HA, hippuric acid; IAA, indole-3-acetate; IxS, indoxyl sulfate; KA, kynurenic acid; Kyn, kynurenine; PAGln, phenylacetylglutamine; pCG, p-cresyl glucuronide; pCS, p-cresyl sulfate; PhG, phenyl glucuronide; Phe, phenylalanine; PhS, phenyl sulfate; TMAO, trimethylamine N-oxide; Trp, tryptophan; Tyr, tyrosine.

**Supplementary Table 5: Associations between symptom variables and metabolites of interest.** Uncorrected linear regression analysis of associations between symptom variables and metabolites, adjusted by Benjamini-Hochberg correction. Effect sizes (partial eta squared values, η_p_^2^), *P* values and *q_FDR_*values as indicated.

**Supplementary Table 6: Associations between symptom variables and metabolites of interest with covariate correction.** Linear regression analysis of associations between symptom variables and metabolites, controlling for dietary protein (IAA, Kyn) or fiber (propionate) and PPI intake, adjusted by Benjamini-Hochberg correction. Effect sizes (partial eta squared values, η_p_^2^), *P* values and *q_FDR_* values as indicated.

**Supplementary Table 7: Multivariate analysis of the combined effects of metabolites of interest on symptom variables.** Multivariate models assessed the combined effect of urinary IAA, serum Kyn and plasma propionate on symptoms while controlling for protein and fiber intake, reporting partial eta squared effect sizes (η_p_^2^), *P* values and *q_FDR_* values.

**Supplementary Table 8: Linear models of Individual and combined effects of groups and psychological symptom variables on metabolites of interest.** Linear regression analyses of associations between metabolites of interest with the grouping variable (FD vs. HC) alone, the grouping variable adjusted for symptom variables, or symptom variables alone. All models are controlled for dietary protein (IAA, Kyn) or fiber (propionate) and PPI intake. Effect sizes of main effects of interest (partial eta squared values, η_p_^2^), *P* values and *q_FDR_* values as indicated.

**Supplementary Table 9: Forty-four brain regions of interest from the salience, emotional-arousal and central autonomic networks from the Johns Hopkins University atlas.** Forty-four regions of interest (ROI) from the salience, emotional-arousal and central autonomic networks from the Johns Hopkins University atlas (PMID: 19385016), imported into CONN, were selected for ROI-to-ROI connectivity analysis based on 1) their hypothesized key role in disorders of gut-brain interaction according to a recent Rome working team report, 2) previous resting-state functional magnetic resonance imaging studies in FD, including systematic reviews and meta-analyses.

**Supplementary Table 10: Multivariate generalized linear model analysis of rs-fMRI-derived ROI-to-ROI connectivity matrix.** Multivariate general linear model (GLM) analyses of a region-of-interest (ROI)-to-ROI connectivity matrix incorporating 44 a priori selected ROIs involved in salience, emotional-arousal and central autonomic networks.

**Supplementary Table 11: Multivariate and univariate analysis of variance on rs-fMRI-derived graph-theoretical measures.** Graph theoretical analysis as a network-based approach included comparison of graph measures between FD and HC using a series of one-way ANOVAs (for non-density-dependent graph measures) and MANOVAs with density as repeatedly measured factor and the main effect of group (across densities) as the effect of interest for density-dependent graph measures. The entire set of 180 resulting *P* values was corrected for multiple testing using positive false discovery rate (*q_FDR_*) calculation.

**Supplementary Table 12: Model metrics of between-group partial least squares discriminant analysis of rs-fMRI-derived graph-theoretical measures.** Model metrics of all 540 resting-state functional magnetic resonance imaging-derived graph-theoretical measures (assortativity (global), betweenness centrality (nodal), efficiency (global and nodal), characteristic path length (global and nodal), and clustering coefficient (global and nodal)) included as X-variables in partial least squares discriminant analysis (PLS-DA). Five-fold cross-validation suggested a 1-factor solution with 55% of X-variables (296/540) exceeding the variable importance plot (VIP) threshold of 0.8.

**Supplementary Table 13: Model metrics of partial least squares regression between rs-fMRI-derived graph-theoretical measures and urinary IAA.** Model metrics of all 540 resting-state functional magnetic resonance imaging-derived graph-theoretical measures (assortativity (global), betweenness centrality (nodal), efficiency (global and nodal), characteristic path length (global and nodal), and clustering coefficient (global and nodal)) included as X-variables in partial least squares (PLS) regression analysis. Five-fold cross-validation suggested a 2-factor solution with 59% of X-variables (320/540) exceeding the variable importance plot (VIP) threshold of 0.8.

**Supplementary Table 14: Non-parametric differential fecal genera abundance analysis.** Non-parametric differential absolute genera abundance analysis between FD and HC was performed after application of a prevalence cut-off of ≥ 20% across the samples, by unpaired Mann-Whitney-U tests with Benjamini-Hochberg correction (*q_FDR_*). Effect sizes are reported as rank biserial correlation coefficients (r).

**Supplementary Table 15: Univariate negative binomial differential fecal genera abundance analysis.** Univariate differential absolute genera abundance analysis (≥ 20% prevalence cut-off) between FD and HC, accounting for spectific distribution of absolute microbial cell counts, by negative binomial models with inclusion of a zero-inflated term for genera that followed a zero-inflated distribution. Parameter estimates, standard error (β ± SE), *P* values and Benjamini-Hochberg corrected *q_FDR_* values as indicated.

**Supplementary Table 16: Multivariate negative binomial differential fecal genera abundance analysis corrected for PPI intake and stool consistency.** Multivariate differential absolute genera abundance analysis (≥ 20% prevalence cut-off) between FD and HC, accounting for specific distribution of absolute microbial cell counts, by negative binomial models with inclusion of a zero-inflated term for genera that followed a zero-inflated distribution. In addition to the grouping variable, ‘PPI intake’ (yes or no, nested within ‘group’) and ‘BSS category’ (hard, normal, loose stools) were included as additional independent variables. Parameter estimates, standard error (β ± SE), *P* values and Benjamini-Hochberg corrected *q_FDR_* values as indicated.

**Supplementary Table 17: Permutational analysis of variance between fecal microbiota compositional variation with dysregulated metabolites and symptom variables.** Associations between dysregulated symptom variables or metabolites in FD and fecal microbial composition were analyzed with permutational analysis of variance (PERMANOVA) as described before. Bristol stool scale (BSS) was included in all models as a covariate. Explained proportion of variance (R^2^), *P* values and Benjamini-Hochberg-adjusted *q_FDR_* as indicated.

**Supplementary Table 18: Correlation analysis of differentially abundant fecal genera with dysregulated metabolites and symptom variables.** Correlations between differentially abundant fecal genera (FD vs. HC, ≥ 20% prevalence cut-off) with dysregulated metabolites and symptom variables were reported as Spearman coefficients (r). P values and Benjamini-Hochberg-adjusted *q_FDR_* values as indicated.

**Supplementary Table 19: Correlation analysis of twenty most abundant fecal genera with dysregulated metabolites and symptom variables.** Correlations between twenty most abundant fecal genera with dysregulated metabolites and symptom variables were reported as Spearman coefficients (r). *P* values and Benjamini-Hochberg-adjusted *q_FDR_* values as indicated.

## Acknowledgements

The authors thank all the patients and volunteers for their participation in this study. We thank the personnel of the Endoscopy and Motility unit at the University Hospitals Leuven campus Gasthuisberg for their assistance with clinical study visits, in particular Karlien Geboers and Lieselot Holvoet. The authors thank Leen Rymenans, Chloë Verspecht and Thi Thuy Duyen Nguyen for their contribution to sample analysis. Icons in Figure 2B-C and Supplementary Figure 1A-B were exported with license to publish from BioRender.com.

## Funding

M.C. was supported by a postdoctoral fellowship of the Belgian American Educational Foundation (BAEF). L.W. was supported by Flanders Research Foundation (FWO Vlaanderen) through a doctoral fellowship (1190619N). N.W. is a postdoctoral fellow supported by a KU Leuven C2 grant (C2M/25/041). R.Y.T. and B.D are funded by a postdoctoral fellowship from the Research Fund–Flanders. A.V.V. is a postdoctoral researcher supported by a Marie-Curie grant (HORIZON-MSCA-2023). J.T. is supported by a KU Leuven Methusalem grant (EZX-C9725-METH/14/05). L.V.O. is a research professor of the KU Leuven Special Research Fund. T.V. is a senior clinical researcher supported by the Research Fund-Flanders (FWO, 1830517N), an FWO research grant (G059822N) and a grant from the Clinical Research Fund UZ Leuven (KOOR project financiering). The Raes lab is supported by KU Leuven, VIB and the Rega Institute. This study was supported by a grant of the Biocodex Microbiota Foundation (BMF Belux 2018) and a grant of the clinical research fund (KOOR) of the University Hospitals Leuven. The funder did not influence the results/outcomes of the study despite author affiliations with the funder.

## Data availability

The raw data generated and analyzed during the current study are available from the corresponding author upon reasonable request.

## Notes

**Conflict of interest statement:** The authors disclose no conflicts.

### Competing Interest Statement

The authors have declared no competing interest.

### Author Declarations

Ethics Committee for Research at University Hospitals / Katholieke Universiteit Leuven gave ethical approval for this work (S60953, S60984)

### Summary of Updates

Formatting updated, discussion updated, figure references corrected.

