## Supplementary methods and results for "Tryptophan metabolites are associated with gut-brain alterations in functional dyspepsia"

by Ceulemans *et al*.

**Supplementary methods**

*Study participants*

Patients with FD were men or women between 18 and 64 years old who fulfilled the Rome IV criteria for FD^1^. Exclusion criteria were the presence of an active psychiatric, inflammatory or metabolic disorder. Structural alterations of the upper GI tract were excluded during upper GI endoscopy. Routine histological analysis of duodenal biopsies confirmed the normal appearance of the duodenal mucosa. All patients had a documented gastric biopsy to rule out *Helicobacter pylori* infection. Patient’s medication intake was recorded and PPI intake at the time of sampling was defined as at least one month of one or more daily doses of any PPI before sampling. Healthy volunteers – men or women between 18 and 64 without any GI, psychiatric, inflammatory or metabolic disorder – were recruited as controls through advertisement. Participants did not use antibiotics or immunosuppressants within three months before sampling. Participants refrained from smoking and alcohol use within two days before sampling.

*Evaluation of symptoms and macronutrient intake*

Gastrointestinal symptoms were assessed by the Patient Assessment of Upper GI Symptom Severity (PAGI-SYM)^2^ and Leuven Postprandial Distress Scale (LPDS)^3^. Extraintestinal and psychological symptoms were assessed by the Generalized Anxiety Disorder-7 questionnaire (GAD7)^4^, Patient Health Questionnaire-9 (PHQ9)^5^, Perceived Stress Scale (PSS)^6^, Visceral Sensitivity Index (VSI)^7^, Post-Traumatic Stress Disorder (PTSD)^8^, Childhood Trauma Questionnaire (CTQ)^9^, Positive and Negative Affect Schedule (PANAS)^10^, and the shortened PHQ15 questionnaire without GI-specific symptoms to evaluate extraintestinal symptom reporting (PHQ12)^11^. Macronutrient intake was estimated through a validated online food frequency questionnaire (FFQ), reporting average daily total energy (kCal/day), protein (g/day), carbohydrate (g/day), fiber (g/day), fat (g/day) and water (mL/day) intake^12^.

*Targeted metabolite analyses*

A validated panel of aromatic amino acids (AAA), host and microbial AAA metabolites, and uremic toxins (**Figure 2A**) was quantified via ultra-performance liquid chromatography-tandem mass spectrometry as previously published^13^ on spot urine samples and fasted serum samples. Creatinine was measured in urine samples via routine enzymatic assay, and urinary metabolite concentrations were normalized to creatinine levels to mitigate dilution effects. Measurement of SCFA was performed as previously described following optimized gas chromatography-mass spectrometry protocols for frozen fecal^14^ and fasted plasma samples^15^.

*Fecal microbiota analysis*

Quantitative microbiota profiling was performed by sequencing the V4 region of the 16S rRNA gene using the 515F (GTGYCAGCMGCCGCGGTAA) and 806R (GGACTACNVGGGTWTCTAAT) primer pair on the Illumina MiSeq platform (MiSeq Reagent Kit v2, 500 cycles, 15.38 % PhiX, 2 × 250 paired-end reads) at the VIB Nucleomics core laboratory (Leuven, Belgium) as described before^16, 17^. All fecal samples were processed in batch at the end of the study. The microbial load of frozen fecal samples was determined by flow cytometry on diluted fecal suspensions as detailed previously^18^. Microbial counts were determined using an Accuri C6 flow cytometer coupled to Accuri CSampler software (BD) with constant instrument and gating settings for all samples, as previously described^16, 18^. R (v4.4.1) was used for bioinformatics analyses, applying the LotuS^19^ and DADA2 (v1.6) pipelines^20^. The QMP matrix was built as described previously^18^. In brief, samples were rarefied to an even sampling depth measured as reads per cell as an integral part of the established QMP pipeline, defined as the ratio between sampling size (16S rRNA gene copy number-corrected sequencing depth) and microbial load per sample (cell count per gram of frozen fecal material). A minimum rarefied read count of 150 was used and rarefied genus abundances were converted into numbers of cells per gram. Taxonomy was assigned up to the genus level, while unassigned amplicon sequence variants (ASVs) at any taxonomic level were labeled with the prefix ‘uc’ to avoid a lack of labels.

*Functional magnetic resonance imaging*

*Preprocessing*

Results included in this manuscript come from preprocessing performed using *fMRIPrep* 20.0.6 (RRID:SCR_016216)^21^, which is based on *Nipype* 1.4.2 (RRID:SCR_002502)^22^.

*Anatomical data preprocessing*

The T1-weighted (T1w) image was corrected for intensity non-uniformity (INU) with N4BiasFieldCorrection^23^, distributed with ANTs 2.2.0 (RRID:SCR_004757)^24^, and used as T1w-reference throughout the workflow. The T1w-reference was then skull-stripped with a *Nipype* implementation of the antsBrainExtraction.sh workflow (from ANTs), using OASIS30ANTs as target template. Brain tissue segmentation of cerebrospinal fluid (CSF), white-matter (WM) and gray-matter (GM) was performed on the brain-extracted T1w using fast (FSL 5.0.9, RRID:SCR_002823)^25^. Volume-based spatial normalization to one standard space (MNI152NLin2009cAsym) was performed through nonlinear registration with antsRegistration (ANTs 2.2.0), using brain-extracted versions of both T1w reference and the T1w template. The following template was selected for spatial normalization: *ICBM 152 Nonlinear Asymmetrical template version 2009c* (RRID:SCR_008796; TemplateFlow ID: MNI152NLin2009cAsym)^26^.

*Functional data preprocessing*

For each of the 1 BOLD runs found per subject (across all tasks and sessions), the following preprocessing was performed. First, a reference volume and its skull-stripped version were generated using a custom methodology of *fMRIPrep*. Susceptibility distortion correction (SDC) was omitted. The BOLD reference was then co-registered to the T1w reference using flirt (FSL 5.0.9)^27^ with the boundary-based registration^28^ cost-function. Co-registration was configured with nine degrees of freedom to account for distortions remaining in the BOLD reference. Head-motion parameters with respect to the BOLD reference (transformation matrices, and six corresponding rotation and translation parameters) are estimated before any spatiotemporal filtering using mcflirt (FSL 5.0.9)^29^. BOLD runs were slice-time corrected using 3dTshift from AFNI 20160207 (RRID:SCR_005927)^30^. The BOLD time-series (including slice-timing correction when applied) were resampled onto their original, native space by applying the transforms to correct for head-motion. These resampled BOLD time-series will be referred to as *preprocessed BOLD in original space*, or just *preprocessed BOLD*. The BOLD time-series were resampled into standard space, generating a *preprocessed BOLD run in MNI152NLin2009cAsym space*. First, a reference volume and its skull-stripped version were generated using a custom methodology of *fMRIPrep*. Several confounding time-series were calculated based on the *preprocessed BOLD*: framewise displacement (FD), DVARS and three region-wise global signals. FD and DVARS are calculated for each functional run, both using their implementations in *Nipype* (following the definitions by Power et al.^31^). The three global signals are extracted within the CSF, the WM, and the whole-brain masks. The head-motion estimates calculated in the correction step were also placed within the corresponding confounds file. The confound time series derived from head motion estimates and global signals were expanded with the inclusion of temporal derivatives and quadratic terms for each^32^. Frames that exceeded a threshold of 0.5 mm FD or 3.0 standardised DVARS were annotated as motion outliers. Four participants (two patients, two controls) were excluded due to excess head movement based on the criterion of >10% motion outliers. All resamplings can be performed with *a single interpolation step* by composing all the pertinent transformations (i.e. head-motion transform matrices, susceptibility distortion correction when available, and co-registrations to anatomical and output spaces). Gridded (volumetric) resamplings were performed using antsApplyTransforms (ANTs), configured with Lanczos interpolation to minimize the smoothing effects of other kernels^33^. Non-gridded (surface) resamplings were performed using mri_vol2surf (FreeSurfer).

Many internal operations of *fMRIPrep* use *Nilearn* 0.6.2 (RRID:SCR_001362)^34^, mostly within the functional processing workflow. For more details of the pipeline, see the section corresponding to workflows in *fMRIPrep*’s documentation^35^.

Finally, functional data were smoothed using spatial convolution with a Gaussian kernel of 8 mm full width half maximum (FWHM) using CONN (RRID:SCR_009550)^36^ release 20.b and SPM^37^ (RRID:SCR_007037) release 12.7771.

*Denoising*

Using CONN (RRID:SCR_009550)^36^ release 20.b and SPM^37^ (RRID:SCR_007037) release 12.7771, functional data were denoised using a standard denoising pipeline including the regression of potential confounding effects characterized by white matter timeseries (5 CompCor noise components), CSF timeseries (5 CompCor noise components), motion parameters and their first order derivatives (12 factors), outlier scans (below 37 factors), session effects and their first order derivatives (2 factors), and linear trends (2 factors) within each functional run, followed by bandpass frequency filtering of the BOLD timeseries between 0.008 Hz and 0.09 Hz. CompCor noise components within white matter and CSF were estimated by computing the average BOLD signal as well as the largest principal components orthogonal to the BOLD average, motion parameters, and outlier scans within each subject's eroded segmentation masks. From the number of noise terms included in this denoising strategy, the effective degrees of freedom of the BOLD signal after denoising were estimated to range from 77.7 to 89.9 (average 88.8) across all subjects.

*ROI-to-ROI connectivity analysis*

*Multivariate GLM*

*ROIs*

44 ROIs from the salience, emotional-arousal and central autonomic networks from the Johns Hopkins University atlas^38^ (**Supplementary Table 7**) (imported into CONN) based on 1) their hypothesized key role in DGBI according to a recent Rome working team report^39^, 2) previous resting-state fMRI studies in FD, including systematic reviews and meta-analyses^40-48^.

*First-level analysis*

ROI-to-ROI connectivity (RRC) matrices were estimated characterizing the functional connectivity between each pair of regions among 44 ROIs. Functional connectivity strength was represented by Fisher-transformed bivariate correlation coefficients from a general linear model (weighted-GLM^]^), estimated separately for each pair of ROIs, characterizing the association between their BOLD signal timeseries. To compensate for possible transient magnetization effects at the beginning of each run, individual scans were weighted by a step function convolved with an SPM canonical hemodynamic response function and rectified.

*Second-level analyses*

Group-level analyses were performed using a General Linear Model (GLM). For each individual connection a separate GLM was estimated, with first-level connectivity measures at this connection as dependent variables (one independent sample per subject), and group as independent variable. Connection-level hypotheses were evaluated using multivariate parametric statistics with random-effects across subjects and sample covariance estimation across multiple measurements. Inferences were performed at the level of individual ROIs. ROI-level inferences were based on parametric multivariate statistics, combining the connection-level statistics across all connections from each individual ROI. Results were thresholded using a familywise corrected p-FWE < 0.05 ROI-level threshold (ROI mass/intensity).

*Graph Theoretical Analysis*

In graph theoretical analysis, the brain is modeled as a network in which nodes represent brain regions and edges denote functional connections, such as correlations or coherence between regional signals. The following complementary metrics were computed to characterize network topology.

Assortativity (global) describes whether brain regions tend to connect to others with a similar number of connections. Positive assortativity indicates that highly connected hubs preferentially connect to other hubs, forming a “rich-club–like” organization, whereas negative assortativity reflects connections between hubs and weakly connected regions. This measure reflects the organizational principle of the brain network and its robustness to damage.

Betweenness centrality (nodal) quantifies how often a brain region lies on the shortest communication paths between other regions. Nodes with high betweenness act as bridges or bottlenecks for information flow and are often critical for integrating information across systems, making them particularly susceptible to dysfunction.

Efficiency reflects the ease of information transfer within the network. Global efficiency measures how efficiently information can be exchanged across the whole network, with higher values indicating shorter average communication paths and greater integrative capacity. Nodal efficiency assesses how efficiently a specific region communicates with all other regions, with high values indicating well-connected regions that facilitate rapid information access.

Characteristic path length represents the average number of steps required to connect any two brain regions via the shortest paths. Shorter global path length indicates more direct communication across the network and is inversely related to global efficiency. At the nodal level, shorter path length signifies central embedding within the network.

Clustering coefficient captures local interconnectivity. The global clustering coefficient measures the overall tendency of regions to form locally interconnected groups, reflecting functional segregation and specialization, whereas nodal clustering coefficient quantifies how strongly a given region’s neighbors are interconnected, indicating participation in tightly knit local circuits.

Together, these graph measures provide complementary information on functional brain organization: efficiency and path length reflect global integration; clustering reflects local segregation; betweenness captures control and mediation; and assortativity indicates network organization and robustness.

Graph Theoretical Analysis were performed using the GraphVar Toolbox^49, 50^ version 2.03a. First, functional connectivity networks (i.e. ROI-to-ROI connectivity matrices) were constructed using the Group Sparse Representation method^51^, a multi-region rather than pairwise-based network construction method shown to suppress spurious correlation and developed to improve performance in classification models, as implemented in the BrainNetClass toolbox^52^ and built into GraphVar. These raw matrices served as inputs (relative thresholding at 0.3, 0.35, 0.4, 0.45, and 0.5 density thresholds representing the 30, 35%, 40%, 45%, and 50% strongest connections, to test robustness of findings across different thresholds; threshold values based on lower thresholds resulting in fragmented networks based on fragmentation check in GraphVar) to calculate the following undirected global graph measures: assortativity (global), betweenness centrality (nodal), efficiency (global and nodal), characteristic path length (global and nodal), and clustering coefficient (global and nodal).

Graph measures were compared between FD patients and HC using a series of one-way ANOVAs (for non-density-dependent graph measures) and MANOVAs with density as repeatedly measured factor and the main effect of group (across densities) as the effect of interest for density-dependent graph measures in JMP^®^ 18.2.1 software (JMP Statistical Discovery LLC, Cary, NC). The entire set of 180 resulting *P* values (i.e., 2 ANOVAs for the non-density-dependent global graph measures characteristic path length and efficiency, 2 MANOVAs for the density-dependent graph measures assortativity and clustering coefficient, 88 ANOVAs for the non-density dependent nodal graph measures characteristic path length and betweenness centrality (2 graph measures x 44 ROIs) and 88 MANOVAs for the density-dependent nodal graph measures clustering coefficient and efficiency) was corrected for multiple testing using the positive FDR (*q_FDR_*) option implemented in SAS 9.4 (TS1M7) proc multtest^53, 54^.

To test the collective ability of the calculated graph measures to discriminate between FD patients and HC, they were entered as independent variables in Partial Least Squares (PLS) Discriminant Analysis using the nonlinear iterative partial least squares (NIPALS) method implemented in JMP^®^ 18.2.1 software (JMP Statistical Discovery LLC, Cary, NC) as it performs well in cases where the number of X values exceeds the number of observations and where X values may be strongly correlated because it reduces the dimensionality of the X-variables by a more limited number of latent factors. Five-fold cross-validation was used to determine the optimal number of factors and avoid overfitting the data, based on a combination of minimum root mean PRESS and Van der Voet T² statistics as well as maximal values for Q² to reach the optimal trade-off between maximally explained variance and minimal risk of overfitting. We used the Tomek Sampling method^55^ implemented in the JMP Unbalanced Classification add-in to account for the unbalanced nature of the classification problem with a 60/40 HC/FD patient ratio, which indicated the exclusion of 2 HCs from the model.

Similarly, the relationship between the calculated graph measures and selected peripheral metabolites was tested using PLS regression for continuous outcomes. Given the smaller sample size (as not all HC included in the brain imaging analysis have metabolomics data), leave-one-out cross-validation was used in this case rather than k-fold. One-way ANOVA was used to compare the standardized factors of the PLS regression model between groups (FD vs. HC), with calculation of Cohen’s d effect sizes, with the following interpretation for magnitudes of difference: small effect size, d of 0.2; medium, 0.5; large, 0.8.

Mediation analysis was performed using the PROCESS macro (v5.0^56^) implemented in SAS 9.4 (TS1M7) with 5000 bootstrap iterations.

*Functional Connectivity Multivariate Pattern Analysis (fc-MVPA)*

Analyses of fMRI data were performed using CONN (RRID:SCR_009550)^36^ release 20.b and SPM^37^ (RRID:SCR_007037) release 12.7771.

*First-level analysis*

Functional connectivity multivariate pattern analyses (fc-MVPA) were performed to estimate the first 6 eigenpatterns characterizing the principal axes of heterogeneity in functional connectivity across subjects. From these eigenpatterns, 6 associated eigenpattern-score images were derived for each individual subject characterizing their brain-wide functional connectome state. Eigenpatterns and eigenpattern-scores were computed separately for each individual seed voxel in the whole brain as the left- and right- singular vectors, respectively, from a singular value decomposition (group-level SVD) of the matrix of functional connectivity values between each seed voxel and the rest of the brain (a matrix with one row per target voxel, and one column per subject). Individual functional connectivity values were computed from the matrices of bivariate correlation coefficients between the BOLD timeseries from each pair of voxels, estimated using a singular value decomposition of the z-score normalized BOLD signal (subject-level SVD) with 64 components separately for each subject.

*Second-level analyses*

Group-level analyses were performed using a General Linear Model (GLM). For each individual voxel a separate GLM was estimated, with first-level connectivity measures at this voxel as dependent variables (one independent sample per subject), and group as independent variable in an F-test across the 6 estimated eigenpatterns. Voxel-level hypotheses were evaluated using multivariate parametric statistics with random-effects across subjects and sample covariance estimation across multiple measurements. Inferences were performed at the level of individual clusters (groups of contiguous voxels). Cluster-level inferences were based on nonparametric statistics using Threshold Free Cluster Enhancement (TFCE), with 1000 residual-randomization iterations. Results were thresholded using a familywise corrected p-FWE < 0.05 TFCE-score threshold.

*Statistical analyses*

Patient characteristics were compared between groups (FD vs. HC) by unpaired Wilcoxon tests for continuous data and Fisher’s exact tests for categorical data. Relative levels (z-scored) of individual SCFA were summed to analyze total SCFA levels in parallel with individual SCFA levels. Similarly, AAA and their metabolites were z-scored and aggregated per origin (**Figure 2A**). Principal component analysis (PCA) based on Euclidean distance and permutational analysis of variance (PERMANOVA) was performed on scaled (z-scored) individual metabolite data measured in serum or in urine to evaluate global between-group differences in metabolite profiles.

Linear regression models analyzing between-group differences metabolite, microbiota, dietary or symptom data were preceded by testing for normally distributed residuals. If needed, Box-Cox transformation was applied to continuous dependent and independent variables to meet the requirement of normally distributed residuals. PPI intake nested in the grouping variable (FD vs. HC) was included in models analyzing group effects on symptom scores, dietary estimates and metabolite concentrations. Additionally, metabolite concentration models were adjusted for fiber (SCFA) or protein (AAA and AAA metabolites) intake. Effect sizes were calculated per independent variable and reported as partial eta squared values (η_p_^2^) for multivariable analyses, with the following interpretation for magnitudes of difference: small effect size, η_p_^2^ of 0.01; medium, 0.06; large, 0.14. For targeted analyses (dietary, symptom and metabolite data), nominal *P* values were calculated for each main effect, with *P* < 0.05 considered statistically significant and *P* < 0.1 considered a trend.

Between-group (FD vs. HC) differences in metabolite categories (serum or urinary AAA, host AAA metabolites, microbial AAA metabolites) following PCA (*rda* function in R package *vegan*) were analyzed by permutational analysis of variance (PERMANOVA, 10,000 permutations) on the scaled metabolite matrix (all individual metabolites or only microbiota-derived individual metabolites) per sample type (serum or urine) using the *adonis2* function in R package *vegan* with or without adjusting for PPI and protein intake. The explained proportion of variance (R^2^) and *P* values were reported.

Associations between symptoms and metabolites of interest (urinary IAA, serum Kyn and plasma propionate) were first explored in a non-parametric way by calculation of Spearman correlation coefficients and subsequently confirmed by parametric linear regression models with and without controlling for dietary protein (IAA, Kyn) or fiber (propionate) and PPI intake, reporting partial eta squared values (η_p_^2^). Multivariate models were constructed to assess the combined effect of urinary IAA, serum Kyn and plasma propionate on symptoms while controlling for protein and fiber intake, reporting partial eta squared values (η_p_^2^). Spearman correlations between the three metabolites of interest were calculated to assess potential collinearity. All non-parametric and parametric analyses were adjusted by Benjamini-Hochberg correction (*q_FDR_*) to account for multiple testing.

To reduce the complexity of statistical models and to prevent overinterpretation of small differences in stool consistency, Bristol Stool Scale (BSS) score was treated as a categorical variable with three ordinal levels: constipation-like hard stools (BSS 1-2), normal stools (BSS 3-5) and diarrhea-like loose stools (BSS 6-7). The proportion of stool consistency types between groups was compared by Fisher’s exact test.

Associations of significantly different metabolites and symptoms between FD and HC with microbiota compositional variation were evaluated using PERMANOVA, with BSS as a fixed covariate in all models. Benjamini-Hochberg correction was applied to adjust for multiple simultaneous comparisons (20 tests) and presented as nominal p-values (*P*) and false discovery rate-adjusted *q*-values (*q_FDR_*).

Alpha diversity indices (within-sample; observed richness, Shannon index, inverse Simpson index) were analyzed at the genus-level QMP matrix using the *estimate_richness* function in R package *phyloseq*. Beta (between-sample) diversity was analyzed by principal coordinate analysis (PCoA) based on Bray-Curtis dissimilarity at the genus level using the *ordinate* function in R package *phyloseq*. PERMANOVA with 10,000 permutations using the *adonis2* function in R package *vegan* assessed the main effect of group, PPI intake and BSS on compositional variation. Associations between symptom variables or metabolites that differed significantly between FD and HC and fecal microbial composition were analyzed with PERMANOVA as described before. BSS was included in all models as a covariate. The explained proportion of variance (R^2^), *P* values and Benjamini-Hochberg-adjusted *q*-values (*q_FDR_*) were reported.

Differential absolute genera abundance analysis was performed after application of a prevalence cut-off of ≥ 20% across the samples, retaining 68 bacterial genera belonging to 6 phyla. Within this dataset, the percentage of samples with zero-abundant genera ranged from 0% (genus detected in all samples) to 79% (genus detected in 21% of the samples). First, unpaired Mann-Whitney-U tests with Benjamini-Hochberg correction (68 tests, *q_FDR_*) were performed to evaluate between-group differences in absolute genera abundances. Effect sizes are reported as rank biserial correlation coefficients (r). Next, negative binomial models (family = nbinom2 using *glmmTMB* function in R package *glmmTMB*) were constructed to account for the specific distribution of QMP data, with inclusion of a zero-inflated term (zi = ~1) for genera that followed a zero-inflated distribution. First, ‘group’ (FD vs. HC) was included as the only independent variable, followed by models where ‘PPI intake’ nested within ‘group’ and ‘BSS category’ were included as additional independent variables. Benjamini-Hochberg correction was applied per dependent variable to adjust for multiple simultaneous comparisons and presented as nominal *P* values and Benjamini-Hochberg-adjusted *q* values (*q_FDR_*). Parameter estimates and standard error (β ± SE) are reported.

Correlations with microbial taxa were performed including taxa with a prevalence of ≥ 20% across the samples (68 taxa) unless specified otherwise by calculation of Spearman’s correlation coefficient. Benjamini-Hochberg correction was applied for the number of tests equaling the number of Y-variables (symptoms or fecal genera) included.

**Supplementary results**

*High psychological symptom burden in FD*

We confirmed the previously described psychological symptom burden of FD^57^ in this cohort, evidenced by significantly higher symptom scores for anxiety (GAD7; η_p_^2^ = 0.35, *P* < 0.0001), perceived stress (PSS; η_p_^2^ = 0.21, *P* = 0.0004), depression (PHQ9; η_p_^2^ = 0.69, *P* < 0.0001), extraintestinal symptoms (PHQ12; η_p_^2^ = 0.59, *P* < 0.0001), GI-specific anxiety (VSI; η_p_^2^ = 0.70, *P* < 0.0001) and posttraumatic stress (PTSD; η_p_^2^ = 0.28, *P* < 0.0001) in patients with FD in comparison to HC, besides more severe GI-specific symptoms (LPDS; η_p_^2^ = 0.67, *P* < 0.0001 and PAGI-SYM; η_p_^2^ = 0.73, *P* < 0.0001) as anticipated (**Figure 1**). Patients with FD also presented with lower positive affect (PANAS+; η_p_^2^ = 0.22, *P* = 0.0002) and higher negative affect (PANAS-; η_p_^2^ = 0.22, *P* = 0.0001) scores (**Figure 1**). Childhood trauma severity (CTQ) was not elevated in FD (η_p_^2^ = 0.006, *P* = 0.51). PPI intake was not associated with alterations in any of the GI or extraintestinal symptom measures (all *P* > 0.1), reflecting that patients with FD taking PPI were symptomatic, as per inclusion criteria for this study (**Supplementary Table 2**). As reported before^58^, food frequency questionnaires revealed differences in dietary patterns between study groups, with lower estimated intake of fiber (η_p_^2^=0.12, *P*=0.015) and protein (η_p_^2^=0.21, *P=*0.0012) in FD compared to HC. Intake of other macronutrients and total energy intake was similar in FD (all *P>*0.1) (**Supplementary Table 3**).

*The fecal microbiota in FD is similar compared to HC and predominantly shaped by intestinal transit time*

The Bristol Stool Scale (BSS) was used to describe the consistency of each individual stool sample as a proxy of intestinal transit time. The proportion of abnormally hard stools (BSS 1-2) and loose stools (BSS 6-7) was similar between FD and HC (*P* = 0.81) (**Figure 5A**). Fecal microbial load as measured by flow cytometric quantification of bacterial cell counts in fecal suspensions did not differ between groups (η_p_^2^ = 0.008, *P* = 0.45) (**Figure 5B**), although, as expected, it decreased with looser stool consistency (η_p_^2^ = 0.094, *P* = 0.030), independent of PPI intake (η_p_^2^ = 0.005, *P* = 0.54) (**Supplementary** **Figure 3A**). In addition, the variance in microbial counts between groups was similar (F = 1.11, *P* = 0.78), indicating that the individual microbiota of patients with FD were not extremely dense or depleted compared to HC.

Genus-based alpha diversity quantified by the number of observed genera, Shannon index and inverse Simpson index was similar in FD and HC (all *P* > 0.1) (**Figure 5C**), while microbial richness decreased with looser stool consistency (observed genera: η_p_^2^ = 0.13, *P* = 0.0068; Shannon index: η_p_^2^ = 0.077, *P* = 0.058) (**Supplementary** **Figure 3B**). Next, we assessed beta-diversity on the QMP matrix as a measure of compositional difference. Overall, both groups exhibited a similar fecal microbial composition as no separate clustering was observed using principal coordinate analysis (PCoA, Bray-Curtis dissimilarity) (**Figure 5D**). PERMANOVA confirmed the absence of a significant difference between groups (R^2^ = 0.011, *P_PERMANOVA_* = 0.63). Stool consistency explained a significant proportion of compositional variance in this cohort (R^2^ = 0.047, *P_PERMANOVA_* = 0.025) while controlling for study group and PPI intake (**Supplementary** **Figure 3C**), in line with previous reports^16, 59, 60^. Therefore, BSS was included as a covariate in downstream microbial analyses.

Analysis of differentially abundant bacterial taxa between groups was performed at the genus level, retaining 68 bacterial genera belonging to six phyla after applying a prevalence cutoff of ≥ 20%. Initially, non-parametric tests indicated higher absolute fecal *Butyricicoccus* abundance in FD (r = 0.32, *P* = 0.02), although no differentially abundant genera persisted after FDR correction (68 tests, all *q_FDR_* = 0.99) (**Supplementary Table 14**). To better account for the specific distribution of absolute bacterial taxa abundances, we applied negative binomial models with an additional zero-inflated term for taxa that were not detected in all samples. Univariable analyses for between-group differences pointed toward decreased *Odoribacter* (B ± SE -0.79 ± 0.22, *q_FDR_* = 0.023) and *Turicibacter* (-0.90 ± 0.26, *q_FDR_* = 0.024) abundance in FD, with a trend for increased *Flavonifractor* (0.71 ± 0.24, *q_FDR_* = 0.067) abundance (**Figure 5F**, **Supplementary Table 15**). When also including PPI intake and BSS as additional independent variables, similar trends for decreased *Odoribacter* (-0.68 ± 0.24) and *Turicibacter* (-0.81 ± 0.28), and increased *Flavonifractor* (0.77 ± 0.27; all *q_FDR_* = 0.075) persisted, as well as for decreased unclassified *Clostridia* (-1.11 ± 0.37, *q_FDR_* = 0.075) (**Figure 5F**, **Supplementary Table 16**). PPI intake at the time of sampling was linked to significantly higher fecal *Streptococcus* (1.38 ± 0.36) abundance, but lower *Bifidobacterium* (-1.43 ± 0.37; both *q_FDR_* = 0.0042) abundance (**Figure 5F**, **Supplementary Table 16**). Of note, the absolute *Prevotella* abundance was lower in fecal samples with a hard consistency (BSS 1-2), while loose stools (BSS 6-7) were associated with lower unclassified *Veillonellaceae*, *Collinsella*, *Coprococcus*, *Dorea*, *Oscillibacter* and unclassified *Coriobacteriaceae* abundance (**Figure 5F**, **Supplementary Table 16**). In summary, despite an overall similar fecal microbial composition, the quantitative abundance of select taxa was different in patients with FD. These shifts were independent of PPI intake and intestinal transit time, although transit time was linked to a higher number of differentially abundant fecal genera.

*Fecal microbiota community variation is closely associated with peripheral metabolite profiles*

Even though the gut microbiota in FD closely resembled that of HC, we evaluated if the marked differences between study groups in metabolites and symptoms were associated with the composition of the gut microbiota, especially considering the microbial origin of various metabolites included in the current analysis. Therefore, we tested for associations between the significantly different metabolites in FD and HC, as well as symptom variables, using PERMANOVA. BSS was included in all models as a covariate given its significant contribution to microbiota compositional variation, which was confirmed in these models (R^2^_BSS_ range: 0.045 - 0.058; *P_PERMANOVA_* < 0.031; *q_FDR_* < 0.031) (**Supplementary Table 17**). Of all tested symptom variables, only GAD7 tended to be associated with microbiota compositional variation (R^2^ = 0.020, *q_FDR_* = 0.099). In contrast, both the aggregated microbial metabolites in serum (R^2^ = 0.048, *q_FDR_* = 0.047) and urine (R^2^ = 0.024, *q_FDR_* = 0.049), as well as host metabolites in serum (R^2^ = 0.027, *q_FDR_* = 0.025) were significantly associated with microbiota composition. Similar significant associations with microbial community variation were found for total plasma SCFA (R^2^ = 0.031, *q_FDR_* = 0.016) and individual metabolites (plasma propionate: R^2^ = 0.029, *q_FDR_* = 0.022; urinary IAA: R^2^ = 0.023, *q_FDR_* = 0.049; urinary PAGln: R^2^ = 0.034, *q_FDR_* = 0.0090; serum PAGln: R^2^ = 0.038, *q_FDR_* = 0.0049; and serum Kyn: R^2^ = 0.026, *q_FDR_* = 0.032) (**Supplementary Table 17**).

*Specific taxa are associated with metabolite and symptom profiles*

Next, we performed correlation analyses between the four differentially abundant bacterial genera in FD compared to HC with select metabolites and symptoms that were substantially different between the study groups. Unclassified *Clostridia* were significantly and positively associated with various urinary and systemic AAA metabolites, but not with any of the symptom measures (**Supplementary Figure 4A**, **Supplementary Table 18**). In contrast, no associations were found for *Flavonifractor*, *Odoribacter* and *Turicibacter* with any of the selected metabolite levels, nor with the included GI and extraintestinal symptoms (**Supplementary Figure 4A**, **Supplementary Table 18**).

The prevalence of the four differentially abundant genera across all fecal samples in this cohort was 33% (25/76), 57% (43/76), 53% (40/76) and 32% (24/76), respectively. We next asked whether symptom profiles and metabolite levels were linked to any of the most abundant bacterial genera. To this end, we repeated the correlation analyses including the twenty most abundant bacterial genera and the same set of symptom and metabolite variables. Interestingly, a cluster consisting of *Anaerostipes* and *Fusicatenibacter* was strongly associated with a mild symptom profile, suggesting a beneficial role for these genera (**Supplementary Figure 4B**, **Supplementary Table 19**). Reduced plasma propionate and total SCFA levels in FD tended to be associated with a lower fecal abundance of *Collinsella* and *Bifidobacterium* (**Supplementary Figure 4B**, **Supplementary Table 19**). Microbial AAA metabolite concentrations were linked to a separate bacterial cluster containing *Clostridium IV* and unclustered *Ruminococcaceae*, while the association with *Clostridium IV* appeared to be driven by PAGln, both in serum and urine (**Supplementary Figure 4B**, **Supplementary Table 19**). The differential abundance of host-derived AAA metabolites, as well as serum Kyn and Phe was not linked with specific fecal bacterial abundance, likely owing to their non-microbial origin (**Supplementary Figure 4B**, **Supplementary Table 19**). Urinary IAA levels tended to correlate positively with fecal *Prevotella* abundance (ρ = 0.34, *q_FDR_* = 0.092), but inversely with *Bacteroides* abundance (ρ = -0.31, *q_FDR_* = 0.092) (**Supplementary Figure 4B**, **Supplementary Table 19**). We could not reliably test the previously reported association of IAA with *Lactobacillus* abundance^61^ as this genus was only detected in less than 20% of the fecal samples. In summary, non-overlapping clusters of specific bacteria correlate separately with psychological symptoms and microbial metabolites. This suggests that the observed associations between select intestinal metabolites and the combined GI and extraintestinal symptom profile in FD do not share a mutual gut microbiota signature.

*Resting-state functional connectivity in salience, emotional-arousal, and central autonomic brain networks is altered in FD*

In addition to the differential pairwise connectivity relationships between groups using functional connectivity multivariate pattern analysis (fc-MVPA), we applied graph theoretical analysis as a network-based approach and compared graph measures between FD and HC (FDR-corrected for 180 tests). Three out of the four global graph measures were significantly different between FD patients and HC ((M)ANOVA), with higher characteristic path length in HC, and higher efficiency and clustering coefficient in FD (**Supplementary Table 11**). For between-group analysis in nodal graph measures using (M)ANOVA, significant differences were found for characteristic path length in 30/44 ROIs, for betweenness centrality in 3/44 ROIs, for clustering coefficient in 20/44 ROIs, and for efficiency in 27/44 ROIs (**Supplementary Table 11**).

**Supplementary Figure legends**

**Supplementary Figure 1:** **Patients with FD exhibit a distinct serum microbial metabolite profile.** Principal component analysis of (**A**) serum microbial AAA metabolites, and (**B**) urinary AAA and AAA metabolites. Permutational analysis of variance (PERMANOVA, 10,000 permutations) quantified explained proportion of variance (R^2^) by ‘Group’.

**Supplementary Figure 2:** **IAA, propionate and Kyn are associated with gastrointestinal and comorbid psychological symptoms in FD.** Linear relationships between psychological (GAD7, PSQ9, PHQ12) and gastrointestinal (LPDS) symptom scores with (**B**) urinary indole-3-acetate (IAA), (**C**) plasma propionate, and (**D**) serum kynurenine (Kyn).

**Supplementary Figure 3:** **Stool consistency is associated with fecal microbiota composition.** Differences in fecal microbial (**A**) cell counts and (**B**) alpha diversity indices (genus-level) by stool sample consistency. Linear regression with main ‘Group’ effect and ‘PPI intake’ nested in the grouping variable or main ‘BSS category’ effect. *P* values represent main ‘BSS category’ effects. (**C**) Principal coordinate analysis (PCoA, Bray-Curtis) of fecal microbiota samples by stool sample consistency (Bristol Stool Scale, BSS). Permutational analysis of variance (PERMANOVA, 10,000 permutations) quantified explained proportion of variance (R^2^) by ‘BSS category’.

**Supplementary Figure 4: Specific taxa are associated with metabolite and symptom profiles.** Heatmap of associations between altered symptom scores and metabolites in FD with (**A**) differentially abundant fecal genera between FD and HC, and (**B**) twenty most abundant fecal genera. Color scale depicts Spearman correlation coefficients. Benjamini-Hochberg adjusted significance depicted as open (*q_FDR_* < 0.1) or filled circles (*q_FDR_* < 0.05).

**Supplementary Figure 5:** **Resting-state functional brain connectivity accurately predicts urinary IAA levels.** Factor loading plots for (**A**) partial least squares discriminant analysis (PLS-DA) and (**B**) PLS regression analysis using functional connectivity-based graph measures (X-variables). Plots depict Y variables in blue (**A**: FD and HC, **B**: urinary IAA), individual FD (black circles) and HC (black triangles) subject loadings, and X-variables (green dots, 540 resting-state functional magnetic resonance imaging (fMRI)-based graph measures) to enter the final PLS-DA and PLS regression models, respectively. (**C**) Linear relationship between observed urinary IAA levels and predicted urinary IAA levels by PLS regression. Pearson correlation coefficient (r) and *P* value as indicated. (**D**) Leave-one-out cross-validation procedure outcomes for urinary IAA, plasma propionate and serum kynurenine (Kyn) suggested a 2-factor solution based on a minimum value for the root mean PRESS statistic for the null model for urinary IAA, while 0 factors were suggested for propionate and Kyn. (**E**) Between-group differences in standardized factors of the PLS regression model explaining 14.9% and 11.9% variance in the graph measures, respectively. One-way analysis of variance.
