## Supplementary figures and images for "Tryptophan metabolites are associated with gut-brain alterations in functional dyspepsia"

### Supplementary Figure 1

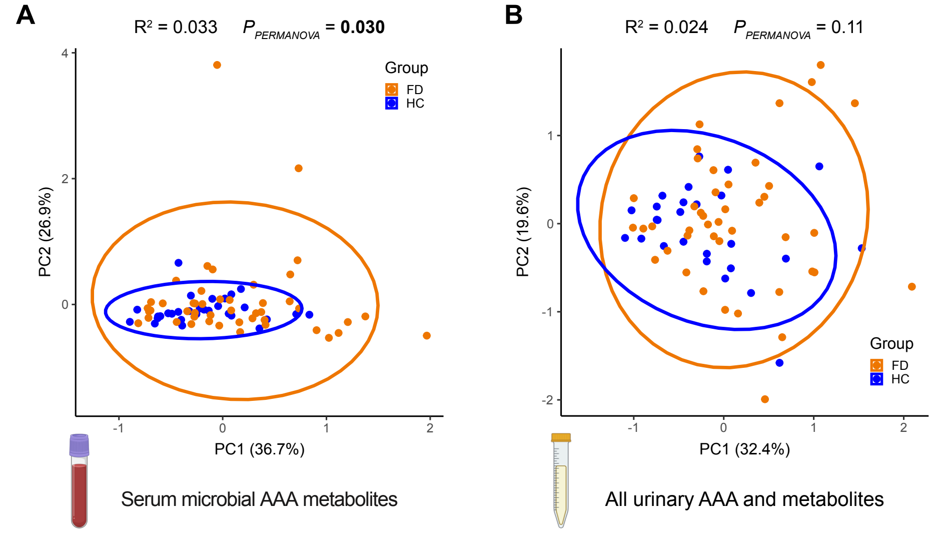


#### Supplementary Figure 1

### Supplementary Figure 2

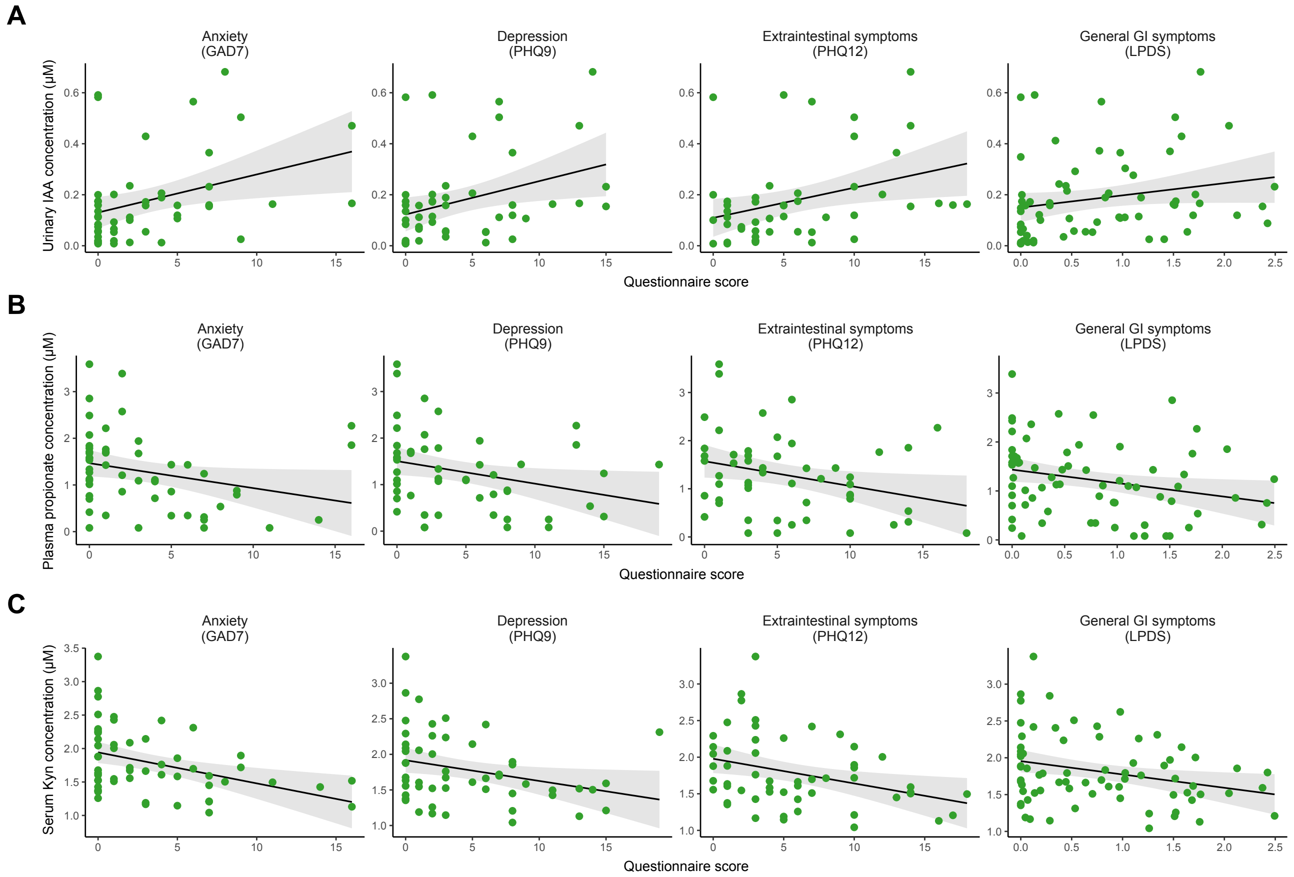


**Supplementary Figure 2**

### Supplementary Figure 3

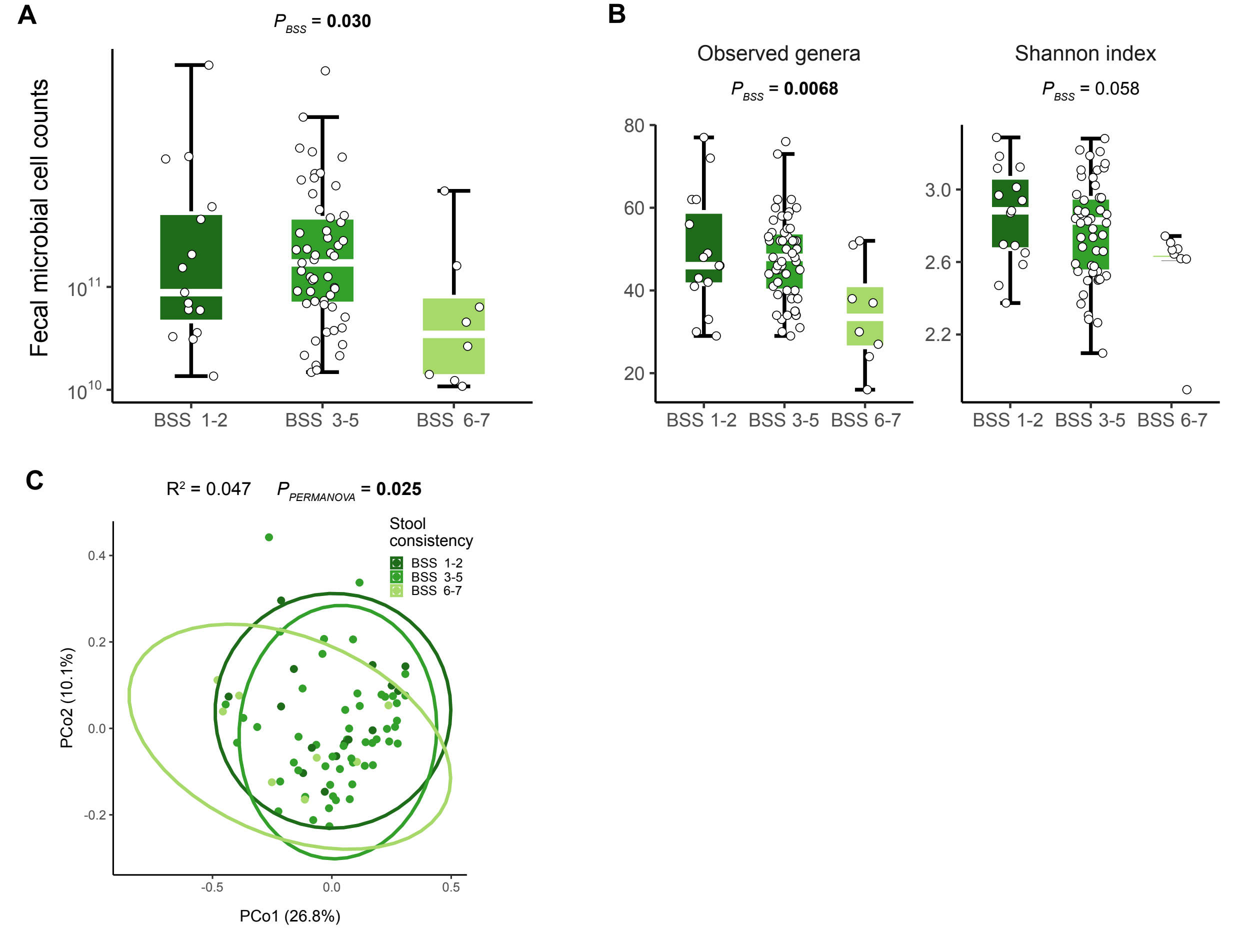


**Supplementary Figure 2**

### Supplementary Figure 4

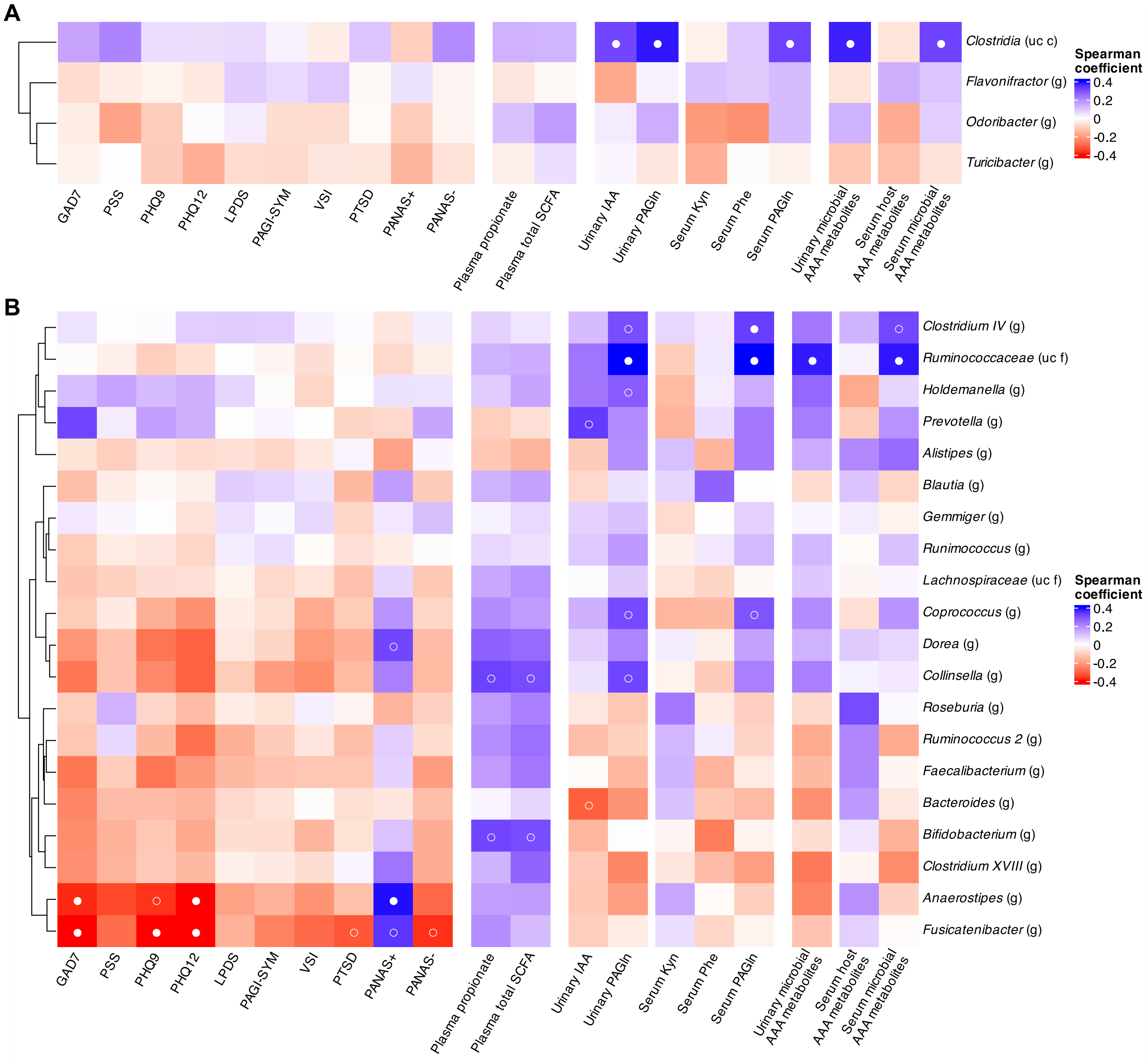


#### Supplementary Figure 3

### Supplementary Figure 5

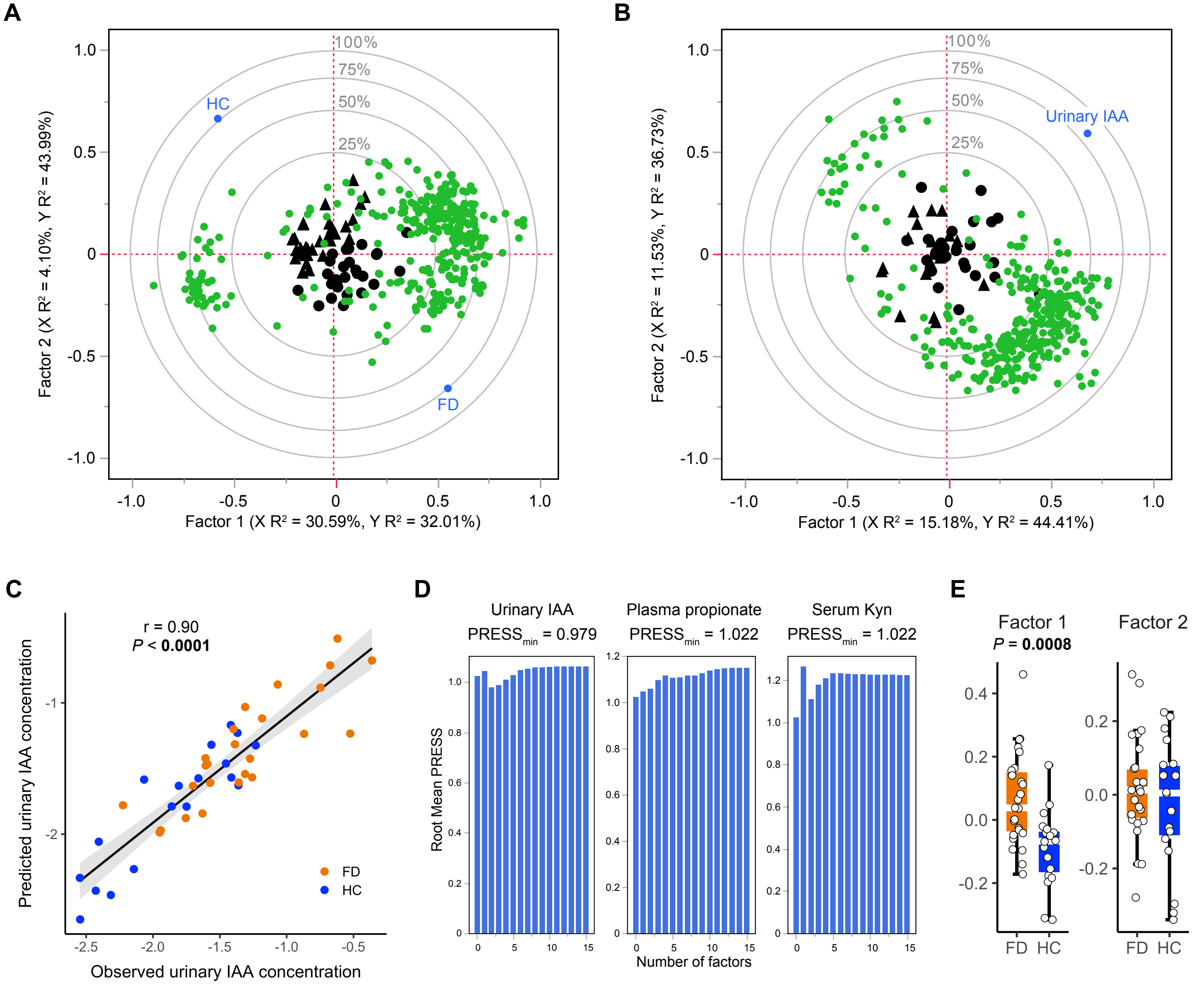


**Supplementary Figure 4**
